## Supplementary material for "Re-assessing thermal response of schistosomiasis transmission risk: evidence for a higher thermal optimum than previously predicted": https://docs.google.com/document/d/1hIcvFzCR42cBX1B-UHI0htxMUXhbiHuh8cx0zqJRedo/edit: Thermal sensitive model of schisto, Supplementary material .pdf (from another version of this preprint: version 1, 2024-01-07)

### The supplementary material of study: "re-assessing thermal response of schistosomiasis transmission risk: evidence for a higher thermal optimum than previously predicted"

#### Contents

The mechanistic model ... 2

Parameters ... 4

Constant parameters ... 4

The factor of density-dependent fecundity rate of snails ... 4

The reduction factor of the fecundity rate due to infection ... 4

The maximum rate of snail invasion ... 4

Constant human population having schistosomiasis ... 4

The number of eggs produced by a mature worm ... 5

The maturation rate of the worms in the human body ... 5

The mortality rate of worms due to human death ... 5

The mortality rate of worms ... 5

Nonconstant parameters ... 5

The fecundity rate of snails ... 7

The mortality rate of snails ... 14

The prepatent period in snails ... 22

The number of cercarial released per snail ... 31

The mortality rate of miracidia ... 34

The mortality rate of cercaria ... 38

The mortality rate of infected snails ... 41

The transmission rate of schistosomiasis in snails ... 48

The transmission rate of schistosomiasis in humans ... 55

Basic reproduction number ... 60

The calculation of basic reproduction number ... 60

Model comparison with GNTD data ... 62

GNTD prevalence data and temperature ... 62

### The mechanistic model

$$\frac{dS}{dt} = (\nu_s(T) - (S + P + I)\nu)(S + rP) - \lambda(T)S - \mu(T)S$$
$$\frac{dE}{dt} = \lambda(T)S - \left(\sigma_s(T) + \mu_i(T)\right)P$$
$$\frac{dI}{dt} = \sigma_s(T)P - \mu_i(T)I$$
$$\frac{dW}{dt} = C_w\beta_h(T)\frac{\nu_c(T)}{\mu_c(T)}I - \sigma_p W$$
$$\frac{dW_m}{dt} = \sigma_p W - \left(\mu_h + \mu_p\right)W_m$$

where the force of infection

$$\lambda(T) = \Lambda\left(1 - e^{-\beta_s(T)\frac{h\delta_e(T)\nu_e W_m/\mu_m(T)}{S+E+I}}\right)$$

Table 1: The parameters' description

| Paramete r | Description | Unit |
|---|---|---|
| $\beta_s(T)$ | Transmission rate of schistosomiasis in snails | $s/p$ |
| $\beta_h(T)$ | Transmission rate of schistosomiasis in humans | $1/h * d$ |
| $\nu_s(T)$ | Fecundity rate of snails | $1/d$ |
| $\mu(T)$ | Mortality rate of snails | $1/d$ |

| | | |
|---|---|---|
| $\delta_e(T)$ | Probability of hatching success of miracidia | *p/e* |
| $\mu_m(T)$ | Mortality rate of miracidia | *1/d* |
| $\mu_c(T)$ | Mortality rate of cercariae | *1/d* |
| $\sigma_s(T)$ | Prepatent period in snails | *1/d* |
| $\mu_i(T)$ | Mortality rate of infected snails | *1/d* |
| $v_c(T)$ | Number of cercariae released by a snail per day | *p/s*d* |
| $v$ | Factor of density-dependent fecundity rate | *1/s*d* |
| $r$ | Reduction factor of fecundity rate due to infection | - |
| $v_e$ | Number of eggs produced by a mature worm | *e/p*d* |
| $\Lambda$ | Maximum rate of snail invasion | *1/d* |
| $\sigma_p$ | Maturation rate of worms in human body | *1/d* |
| $h$ | Constant human population having worms | $h$ |
| $\mu_p$ | Mortality rate of worms | $1/d$ |
| $\mu_h$ | Mortality rate of worms due to human death | *1/d* |

The explicit format of dynamic consist of seven compartments, we embed into two fast dynamics (miracidia and cercariae life span) into slow dynamic by setting the system into the equilibrium points.

$$\frac{dM}{dt} = h\nu_e\delta_e(T)W_m - \mu_m(T)M = 0 \Rightarrow M^* = \frac{h\nu_e\delta_e(T)W_m}{\mu_m(T)}$$

$$\frac{dC}{dt} = \nu_c(T)I - \mu_c(T)C = 0 \Rightarrow C^* = \frac{\nu_c(T)}{\mu_c(T)}I$$

Note that some portions of miracidia and cercariae are transmitted to snails' tissue and human body and these portions are neglected due to their magnitude compared to mortality rate of miracidia and cercaria.

### Parameters

The mechanistic model has two sets of parameters. The constant parameters which do not vary with temperature and the non-constant parameters which vary with temperature.

#### Constant parameters

In this section, we discuss the parameters values not varying with temperature. The list of these parameters: Factor of density-dependent fecundity rate, Reduction factor of fecundity rate due to infection, Number of eggs produced by a mature worm, Maximum rate of snail invasion, Maturation rate of worms in human body, Constant human population having worms, Mortality rate of worms, Mortality rate of worms due to human death.

#### The factor of density-dependent fecundity rate of snails

The parameter is denoted in the mechanistic model with $\nu$ and has the unit of 1/(snail*day). The parameter represents the rate of contribution to reach the carrying capacity per snail in one day if the DFE is stable. For instance, If we assume the carrying capacity is 10000 in case of DFE is stable then, $\nu = 1/10000$. However, since the endemic equilibrium is stable in most areas of our interest [1], we adjust this value to get a desirable snail population size.

#### The reduction factor of the fecundity rate due to infection

The parameter is denoted in the mechanistic model with $r$ and does not have a unit. It represents the drop in the fecundity rate due to disease in the infected snails. So, the values should vary between [0,1]. There are not many resources, but one the study by [2] takes the fecundity rate of infected snails to little less than half of the susceptible snails, so we can assume it drops the fecundity rate by half. In addition, a study by [3], uses the value $r = 0.5$

#### The maximum rate of snail invasion

The parameter is denoted in the mechanistic model with $\Lambda$ and has a unit of 1/day. It regulates the transmission rate of disease in snails [4]. Thus, we manipulate the value of $\Lambda$ to reach the desirable prevalence in snails.

#### Constant human population having schistosomiasis

The parameter is denoted in the mechanistic model with $h$ and has the unit of human. The parameter represents the constant human population in the territory of interest. This value varies across geography.

##### The number of eggs produced by a mature worm

The parameter is denoted in the mechanistic model with $\nu_e$and has the unit of egg/(parasite*day). The value of the parameter represents the eggs released into the environment by one pair of worms in one day. One study by [5] mentions "*S. mansoni* worm pairs laid approximately 350 eggs per day with no change in the apparent rate of egg-laying between eight and 52 weeks after infection, and approximately one-third of the eggs were passed in the feces". [6] "a mated pair of schistosomes is responsible for 200–300 ~ 350/3 eggs per day". We assume this value does not depend upon temperature because the eggs are produced inside the human body.

##### The maturation rate of the worms in the human body

The parameter is denoted in the mechanistic model with $\sigma_p$ and has a unit of 1/day. The value of the parameter represents the maturation rate of worms per day. The average time for maturation of *S.mansoni* is 50-60 days according to [6]. One other resource by [2]uses $\sigma_p = 0.02$ for the maturation rate of worms in the human body for species *S. haematobium* and *S. mansoni.* We assume this value does not depend upon temperature because worms' maturation is happening inside the human body.

##### The mortality rate of worms due to human death

The parameter is denoted in the mechanistic model with $\mu_h$ and has a unit of 1/day. The value of the parameter represents the death rate of humans per day.

##### The mortality rate of worms

The parameter is denoted in the mechanistic model with $\mu_p$ and has a unit of 1/day. It represents the death rate of worms per day. Since parasites live in the human body and humans are endothermic species, this value does not change with the outside temperature. A study by [7] found the *S. mansoni* worm's life span with a 95% confidence interval to be [5.7-10.5] ~ 8 years. Thus, we assume the average lifespan of a worm in the human body is 8 years.

#### Nonconstant parameters

In this section, we discuss the temperature-varying rates from the experiment results that are found in the literature for different species and then use these rates with the R package called rTPC [8] to find a curve for each temperature-dependent parameter. The package performs 24 thermal-sensitive curves for each temperature-dependent parameter and we pick the curve based on three criteria; i) the smallest Akaike information criteria (AIC) score, ii) simpler regarding the number of parameters and iii) biologically applicable. The curves are performed for *S. mansoni* and *S. haematobium* parasites separately and for *Biomphalaria* and *Bulinus* snails separately. In addition, in order to analyze the uncertainty in our curve estimation, we did bootstrapping resampling and found a 95 percent confidence interval around each curve. The following two tables give the selected models and their details. The rest of the section extensively discusses the details about each selected model.

Table 1: The selected curves for *S. mansoni* and *Biomphalaria* species

| The parameter | The selected model | The ranking of the model | The optimal temp | Bootstrapping method | Representation |
|---|---|---|---|---|---|
| Fecundity rate of snail | Gaussian | 1 | 24 °C | Resampling | $r_{max}e^{-0.5\left(\frac{T-T_{opt}}{a}\right)^2}$ |
| Mortality rate of snail | Spain | 1 | 21 °C | Resampling | $r_0e^{aT}(1-be^{cT})$ |
| Mortality rate of inf. Snail | Flinn | 8 | Increasing function | Residual resampling | $\frac{1}{1+a+bT+cT^2}$ |
| Mortality rate of miracidia | Thomas 2017 | 1 | 13.5 °C | Resampling | $ae^{bT}-(c+de^{eT})$ |
| Mortality rate of cercaria | Spain | 2 | 16.5 °C | Residual resampling | $r_0e^{aT}(1-be^{cT})$ |
| Hatching rate of miracidia | Flinn | 1 | Increasing function | Resampling | $\frac{1}{1+a+bT+cT^2}$ |
| Prepatent period in snail | Gaussian | 1 | Increasing function | Resampling | $r_{max}e^{-0.5\left(\frac{T-T_{opt}}{a}\right)^2}$ |
| Cercaria release rate | Gaussian | 1 | 27.5 °C | Resampling | $r_{max}e^{-0.5\left(\frac{T-T_{opt}}{a}\right)^2}$ |
| The transition rate in snails | Spain | 12 | 33 °C | Resampling | $r_0e^{aT}(1-be^{cT})$ |
| The transition rate in humans | Briere2 | 8 | 24 °C | Resampling | $aT(T-T_{min})(T_{max}-T)^{\frac{1}{b}}$ |

Table 1: The selected curves for *S. haematobium* and *Bulinus* species

| The parameter | The selected model | The ranking of model | The optimal temp | Bootstrapping method | Representation |
|---|---|---|---|---|---|
| Fecundity rate of snail | Johnson Lewin | 14 (simpler) | 27.8 °C | Resampling | |

| | | | | | |
|---|---|---|---|---|---|
| Mortality rate of snail | Quadratic | 8 | 15 °C | Resampling | $a + bT + cT^2$ |
| Mortality rate of inf. Snail | Spain | 3 | Increasing function | Residual resampling | $r_0 e^{aT}(1 - be^{cT})$ |
| Mortality rate of miracidia | Spain | 2 | 22.5 °C | no enough data to do resampling | $r_0 e^{aT}(1 - be^{cT})$ |
| Mortality rate of cercaria | Spain | 2 | 16.5 °C | Residual resampling | $r_0 e^{aT}(1 - be^{cT})$ |
| Hatching rate of miracidia | Flinn | 1 | Increasing function | Resampling | $\frac{1}{1+a+bT+cT^2}$ |
| Prepatent period in snail | Gaussian | 1 | Decaying function | Resampling | $r_{max} e^{-0.5\left(\frac{T-T_{opt}}{a}\right)^2}$ |
| Cercaria release rate | Gaussian | 1 | 27.5 °C | Resampling | $r_{max} e^{-0.5\left(\frac{T-T_{opt}}{a}\right)^2}$ |
| The transition rate in snails | Flinn | 8 | 30.5 °C | Residual resampling | $\frac{1}{1+a+bT+cT^2}$ |
| The transition rate in humans | Briere2 | 8 | 24 °C | Resampling | $aT(T - T_{min})(T_{max} - T)^{\frac{1}{b}}$ |

#### The fecundity rate of snails

The parameter is denoted in the mechanistic model with $\nu_s$ has the unit of 1/day. The value represents reproduction of snails in a day. We use Euler- Lotka equation to approximate the fecundity rate [9].

$$1 = \sum_{a}^{w} \lambda^{-a} l(a) b(a)$$

where $\lambda$ is the growth rate, $l$(a) is the fraction of individuals surviving to age *a,* and *b(a)* is the number of offspring born to an individual of age during the time step. $w$ is the longest time to survive. By utilizing the model above

1= $\sum_a^w (prob.of\ surv.till\ maturity) * (daily\ surv.)^a * 0.5\ (eggs/snail/day)(growth\ rate)^{-(a+age\ of\ maturity)}$

Note that fecundity-mortality = growth rate, then fecundity rate is mortality + growth. We will be using: Age at maturity, hatching rate, Daily Survival. Egg/snail/day given in empirical studies to estimate the fecundity rate of snails.

McCreesh et al. [10], *Biomphalaria sudanica*. In this table, the age of maturity and hatching rates are from El-Hassan [11], for *Biomphalaria alexandrina*.

| Temperature | Age at maturity | Hatching rate | Daily survival rate | Egg/snail/day | Fecundity rate |
|---|---|---|---|---|---|
| 13.4°C | 9999 | 0 | 0.990596 | 0.11714 | 0 |
| 15.7°C | 360 | 89.5 | 0.9831741 | 3.71714 | 0 |
| 16.7°C | 107 | 89.5 | 0.994776 | 6.48286 | 0.0349 |
| 18.9°C | 107 | 89.5 | 0.9929975 | 8.52714 | 0.0369 |
| 20.9°C | 107 | 97.9 | 0.9984211 | 8.63857 | 0.0395 |
| 22.8°C | 107 | 97.9 | 0.9910489 | 7.00000 | 0.0386 |
| 26.7°C | 77 | 98.8 | 0.9926992 | 7.03857 | 0.0499 |
| 28.3°C | 77 | 98.8 | 0.977706 | 5.11429 | 0 |
| 29.5°C | 154 | 97.7 | 0.9610102 | 2.96286 | 0 |
| 32.0°C | 360 | 0 | 0.9409337 | 0.58714 | 0 |

De Kock and Van Eeden [12], *Biomphalaria pfeifferi*. The temperature fluctuates in this study. (This study was not used in the simulation due not constant temperature.)

| Temperature | Age at maturity | Hatching rate | Daily survival rate | Egg/snail/day | Fecundity rate |
|---|---|---|---|---|---|
| 18-28$^{o}$ C | 32 | 98 | 0.99748 | 10.22014 | 0.09774 |
| 20.5-25.5$^{o}$ C | 33 | 99 | 0.99368 | 4.024405 | 0.07891 |
| 23$^{o}$ C | 36 | 99 | 0.99631 | 4.941586 | 0.07686 |

Appleton [13], *Biomphalaria pfeifferi*

| Temperature | Age at maturity | Hatching rate | Daily survival rate | Egg/snail/day | Fecundity rate |
|---|---|---|---|---|---|
| 25$^{o}$ C | 42 | 95.6 | 0.99717 | 7.133363 | 0.07058 |
| 27$^{o}$ C | 35 | 88.8 | 0.98991 | 3.652113 | 0.06824 |
| 29$^{o}$ C | N/A | N/A | N/A | 0 | 0 |

El-Hassan [11], *Biomphalaria alexandrina*

| Temperature | Age at maturity | Hatching rate | Daily survival rate | Egg/snail/day | Fecundity rate |
|---|---|---|---|---|---|
| 15° C | 360 | 89.5 | 0.989 | 0.166 | 0.00683 |
| 20° C | 107 | 97.9 | 0.994 | 0.663 | 0.0229 |
| 25° C | 77 | 98.8 | 0.993 | 0.849 | 0.0309 |
| 30° C | 154 | 97.7 | 0.985 | 0.057 | 0.00824 |
| 35° C | 360 | 0 | 0.962 | 0 | 0 |

Sturrock [14], *Biomphalaria pfeifferi*

| Temperature | Age at maturity | Hatching rate | Daily survival rate | Egg/snail/day | Fecundity rate |
|---|---|---|---|---|---|
| 19° C | 110 | 97.9 | 0.98581 | 1.24541 | 0.02720 |
| 25° C | 40 | 98.9 | 0.98647 | 5.03455 | 0.07378 |
| 30° C | 38 | 97.7 | 0.95977 | 1.990214 | 0.06568 |
| 35° C | N/A | 0 | N/A | 0 | 0 |

R. F. Sturrock and B. M. Sturrock [15] *Biomphalaria glabrata*

| Temperature | Age at maturity | Hatching rate | Daily survival rate | Egg/snail/day | Fecundity rate |
|---|---|---|---|---|---|
| 20° C | 112 | 86 | 0.7876 | 5.714 | 0.05856 |
| 25° C | 55 | 96 | 0.9160 | 21.428 | 0.08750 |
| 30° C | 56 | 95 | 0.8439 | 2.857 | 0.06577 |
| 35° C | NA | 0 | NA | 0 | 0 |

Michelson [16], *Biomphalaria glabrata.* In this table, the age of maturity and hatching rates are from El-Hassan [11], for *Biomphalaria alexandrina*.

| Temperature | Age at maturity | Hatching rate | Daily survival rate | Egg/snail/day | Fecundity rate |
|---|---|---|---|---|---|
| $5^0$ | - | - | | 0 | 0 |
| $15^0$ | - | - | | 0 | 0 |
| $20^0$ | 107 | 97.9 | 0. 994 | 2.5/21 = 0.12 | 0.0132 |
| $25^0$ | 77 | 98.8 | 0.993 | (42.2/20+19.2/15+17.1/10+1.7/6)/4=1.35 | 0.0349 |
| $30^0$ | 154 | 97.7 | 0.985 | (0/5+2.5/10+1.3/15+0)/4 =0.0842 | 0.00969 |
| $35^0$ | 360 | 0 | 0.962 | 0 | 0 |

Shiff and Garnett [17], *Biomphalaria pfeifferi.* In this table, hatching rates are from El-Hassan [11], for *Biomphalaria alexandrina*.

| Temperature | Age at maturity | Hatching rate | Daily survival rate | Egg/snail/day | Fecundity rate |
|---|---|---|---|---|---|
| $18^0$ | 11.5*14 = 161 | 89.5 | 0.998 | 0.33 | 0.0138 |
| $22^0$ | 4.5*14 = 63 | 97.9 | 0.996 | 3.58 | 0.0507 |
| $25^0$ | 4.5*14 = 63 | 98.8 | 0.997 | 0.81 | 0.0349 |
| $27^0$ | 5.5*14 = 77 | 98.8 | 0.995 | 1.521 | 0.036 |

El-Hassan [11], *Bulinus truncatus*

| Temperature | Age at maturity | Hatching rate | Daily survival rate | Egg/snail/day | Fecundity rate |
|---|---|---|---|---|---|
| $15^o$ C | 220 | 90.7 | 0.998 | 0.261 | 0.0103 |
| $20^o$ C | 108 | 100 | 0.999 | 0.968 | 0.025 |

| | | | | | |
|---|---|---|---|---|---|
| $25^{o}$ C | 75 | 93.7 | 0.995 | 1.335 | 0.0348 |
| $30^{o}$ C | 135 | 97.5 | 0.970 | 0.548 | 0.0199 |
| $35^{o}$ C | 250 | 95.2 | 0.908 | 0.102 | 0 |

Kalinda et al. [18], *Bulinus globosus.* In this table, the age of maturity and hatching rates are from El-Hassan [11], for *Bulinus truncatus.*

| Temperature | Age at maturity | Hatching rate | Daily survival rate | Egg/snail/day | Fecundity rate |
|---|---|---|---|---|---|
| 15.5°C | N/A | N/A | 0.998 | 0 | 0 |
| 21.2°C | 108 | 100 | 0.993 | 2.59909 | 0.027 |
| 25.8°C | 75 | 93.7 | 0.989 | 2.766767 | 0.037 |
| 31°C | 135 | 97.5 | 0.979 | 2.08225 | 0.019 |
| 35.5°C | N/A | N/A | 0.955 | 0 | 0 |

Shiff [19], *Bulinus globosus.* In this table, hatching rates are from El-Hassan [11] for *Bulinus truncatus*

| Temperature | Age at maturity | Hatching rate | Daily survival rate | Egg/snail/day | Fecundity rate |
|---|---|---|---|---|---|
| $18^{0}$ | 11.5*14 = 161 | 90.7 | 0.994 | 0.3215143 | 0.0137 |
| $22^{0}$ | 6.5*14 = 63 | 100 | 0.993 | 2.285877 | 0.0447 |
| $25^{0}$ | 4.5*14 = 63 | 93.7 | 0.98 | 3.197019 | 0.0497 |
| $27^{0}$ | 4.5*14 = 63 | 93.7 | 0.98 | 1.159799 | 0.0391 |

Kubirizajournal et al.[20], *Bulinus nyassanus.* In this table, the age of maturity is from El-Hassan [11], for *Bulinus truncatus*.

| Temperature | Age at maturity | Hatching rate | Daily survival rate | Egg/snail/day | Fecundity rate |
|---|---|---|---|---|---|
| $22^{0}$ | 108 | 67.11 | 0.993 | 2.846667 | 0.0297 |

| | | | | | |
|---|---|---|---|---|---|
| $25^0$ | 75 | 88.65 | 0.994 | 3.336667 | 0.0427 |
| $28^0$ | 75 | 88.96 | 0.979 | 4.226667 | 0.0463 |
| $31^0$ | 135 | 91.12 | 0.989 | 3.733333 | 0.0451 |

The top five models for *Biomphalaria* snails

| Model | AIC value | BIC |
|---|---|---|
| Gaussian | -176 | -169 |
| Modifiedgaussian | -174 | -166 |
| Oneill | -174 | -166 |
| Pawar | -174 | -166 |
| Sharpeschoolhugh | -174 | -166 |

Gaussian curve is selected and the expression is

$$r_{max}e^{-0.5\left(\frac{T-T_{opt}}{a}\right)^2}$$

See Lynch and Gabriel [21] for additional information. The fitted curve to *Biomphalaria* data as follow

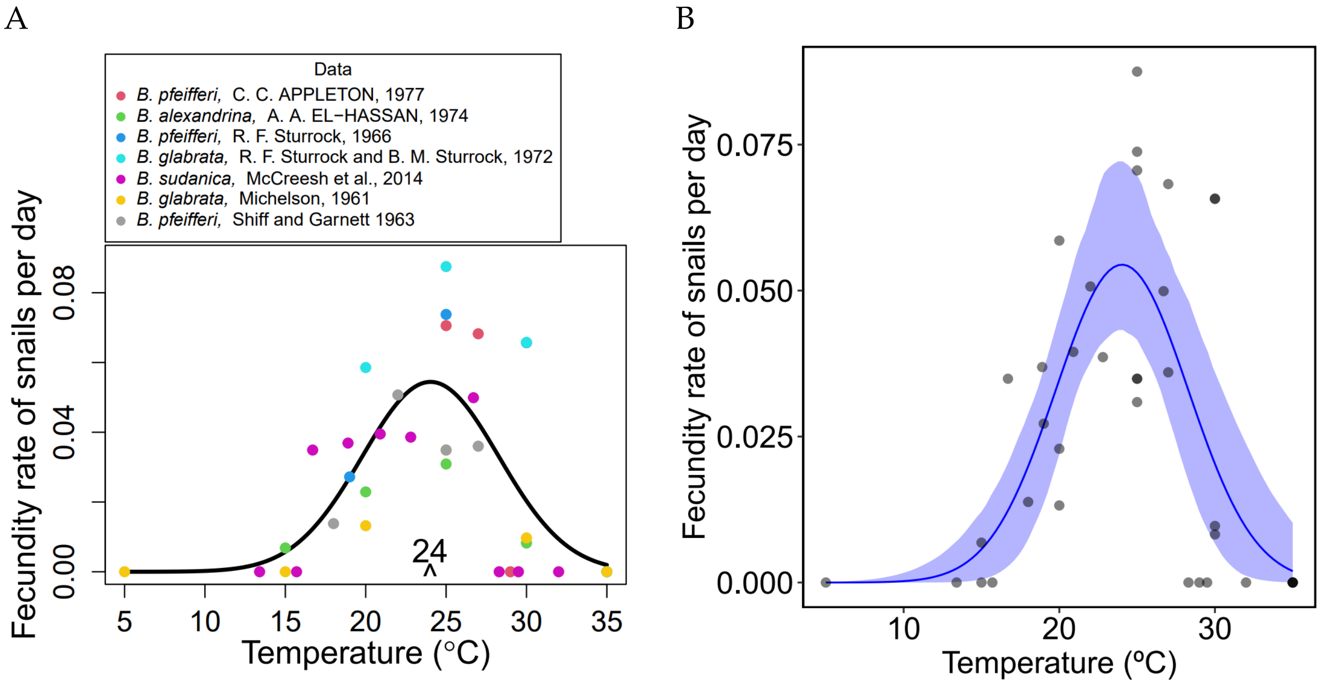

Figure 1: Fecundity rate of snails and confidence intervals. The different colors represent different data resources.

The top five models for *Bulinus* snails

| Model | AIC value | BIC |
|---|---|---|
| Ratkowsky | -134 | -129 |
| Modifiedgaussian | -133 | -128 |
| Oneill | -133 | -128 |
| Gaussian | -132 | -128 |
| Weibull | -132 | -127 |
| Johnson Lewin (14th in ranking) | -128 | -123 |

Johnson Lewin curve is selected and the expression is

$$\frac{r_0 e^{\frac{-e}{k(T+273.15)}}}{1+e^{-\frac{e_h-\left(\frac{e_h}{T_{opt}+273.15}+k ln\left(\frac{e}{e_h-e}\right)\right).(T+273.15)}{k.(T+273.15)}}}$$

See Johnson and Lewin [22] for additional information. The fitted curve to *Bulinus* data as follow

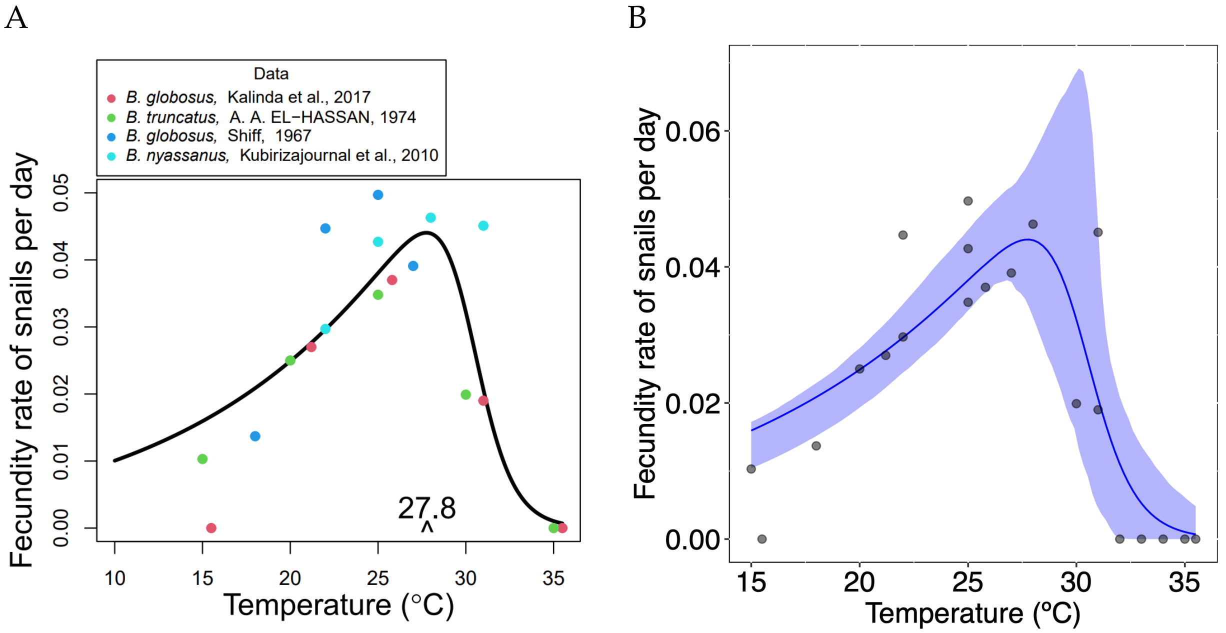

Figure 2: Fecundity rate of snails and confidence interval. The different colors represent different data resources.

#### The mortality rate of snails

The parameter is denoted in the mechanistic model with $\mu$ has the unit of 1/day. The parameter represents the mortality rate of snails per day. We use the exponential growth model

$$\frac{dN(t)}{dt} = -\mu N(t) \Rightarrow \mu = -\frac{log(N/N_0)}{t}$$

with given survival portions of snails each time then take the average of the found rates to get the mortality rate. For instance, given the percentage of survivals $(x_1, \ldots, x_n)$ for times $(t_1, \ldots, t_n)$ at each temperature T. We use the formula $\mu_i = \frac{log(x_i)}{t_i}$ and then take the average of $\mu$ to get the mortality rate for temperature T.

Shiff and Garnett [17], *Biomphalaria pfeifferi.*

| Temperature | Mortality rate |
|---|---|
| $18^0$ | 0.001650303 |
| $22^0$ | 0.002659904 |
| $25^0$ | 0.002373008 |
| $27^0$ | 0.004170385 |

McCreesh et al. [10], *Biomphalaria sudanica*

| Temperature | Mortality rate |
|---|---|
| 13.4°C | 0.009404046 |
| 15.7°C | 0.01682591 |
| 16.7°C | 0.005224019 |
| 18.9°C | 0.007002545 |
| 20.9°C | 0.001578888 |
| 22.8°C | 0.008951098 |
| 26.7°C | 0.007300832 |
| 28.3°C | 0.02229403 |
| 29.5°C | 0.03898983 |
| 32.0°C | 0.05906629 |

Joubert et al. [23], *Biomphalaria pfeifferi*

| Temperature | Mortality rate |
|---|---|
| 34°C | 0.1457006 |
| 36°C | 0.6661235 |

| | |
|---|---|
| 38°C | 1.649401 |
| 40°C | 5.821733 |

De Kock and Van Eeden [12], *Biomphalaria pfeifferi.* Note that here temperature fluctuates.

| Temperature | Mortality rate |
|---|---|
| 23° C, no fluctuation | 0.00369937 |
| 20.5-25.5° C fluctuation | 0.006338469 |
| 18-28° C | 0.002522396 |

Appleton [13], *Biomphalaria pfeifferi*

| Temperature | Mortality rate |
|---|---|
| 25° C | 0.002832593 |
| 27° C | 0.01014083 |
| 29° C | 0.02164846 |

El-Hassan [11], *Biomphalaria alexandrina*

| Temperature | Mortality rate |
|---|---|
| 10° C | 0.01161 |
| 15° C | 0.01077 |
| 20° C | 0.00596 |
| 25° C | 0.00674 |
| 30° C | 0.01505 |

| | |
|---|---|
| 35° C | 0.03891 |
| 37° C | Inf |

Sturrock [14], *Biomphalaria pfeifferi*

| Temperature | Mortality rate |
|---|---|
| 19° C | 0.0142923 |
| 25° C | 0.01361983 |
| 30° C | 0.04106602 |

Foster [24], *Biomphalaria pfeifferi*

| Temperature | Mortality rate |
|---|---|
| 22.85° C | 0.006674626 |
| 24.01° C | 0.01155245 |
| 26.26° C | 0.0260108 |
| 28.07° C | 0.03678792 |

El-Emam and Madsen [25], *Biomphalaria alexandrina*

| Temperature | Mortality rate |
|---|---|
| $10^0$ | NAN |
| $18^0$ | 0.006188814 |
| $26^0$ | 0.006188814 |
| $28^0$ | 0.009373412 |
| $33^0$ | NAN |

El-Emam and Madsen [25], *Bulinus truncatus*

| Temperature | Mortality rate |
|---|---|
| $10^0$ | 0.00256859 |
| $18^0$ | 0.004560943 |
| $26^0$ | 0.003184598 |
| $28^0$ | 0.004560943 |
| $33^0$ | NAN |

Shiff [19], *Bulinus globosus*

| Temperature | Mortality rate |
|---|---|
| $18^0$ | 0.005314696 |
| $22^0$ | 0.006322683 |
| $25^0$ | 0.01045628 |
| $27^0$ | 0.01057945 |

Kalinda et al. [18], *Bulinus globosus*

| Temperature | Mortality rate |
|---|---|
| 15.5°C | 0.00244925 |
| 21.2°C | 0.006901087 |
| 25.8°C | 0.01151302 |
| 31°C | 0.02073716 |
| 35.5°C | 0.04642702 |

Kubirizajournal et al. [20], *Bulinus nyassanus*

| Temperature | Mortality rate |
|---|---|
| $22^0$ | 0.006849573 |
| $25^0$ | 0.00513718 |
| $28^0$ | 0.02054872 |
| $31^0$ | 0.01027436 |

Joubert et al. [23], *Bulinus globosus*

| Temperature | Mortality rate |
|---|---|
| 34°C | 0.02924029 |
| 36°C | 0.1490773 |
| 38°C | 0.8022027 |
| 40°C | 3.540546 |

Joubert et al. [23], *Bulinus africanus*

| Temperature | Mortality rate |
|---|---|
| 34°C | 0.1044946 |
| 36°C | 0.5664857 |
| 38°C | 1.618022 |
| 40°C | 4.344277 |

El-Hassan [11], *Bulinus truncates*

| Temperature | Mortality rate |
|---|---|
| $10^o$ C | 0.00442 |

| | |
|---|---|
| 15° C | 0.00218 |
| 20° C | 0.00144 |
| 25° C | 0.00518 |
| 30° C | 0.03077 |
| 35° C | 0.09622 |
| 37° C | Inf |

The top five models for *Biomphalaria* snails

| Model | AIC value | BIC |
|---|---|---|
| Spain | -223 | -216 |
| Thomas2 | -222 | -213 |
| Joehnk | -219 | -210 |
| Beta | -216 | -207 |
| Ratkowsky | -215 | -208 |

Spain curve is selected and the expression is

$$r_0 e^{aT}(1 - be^{cT})$$

See Cunningham [26] for additional information. The fitted curve to *Biomphalaria* data as follow

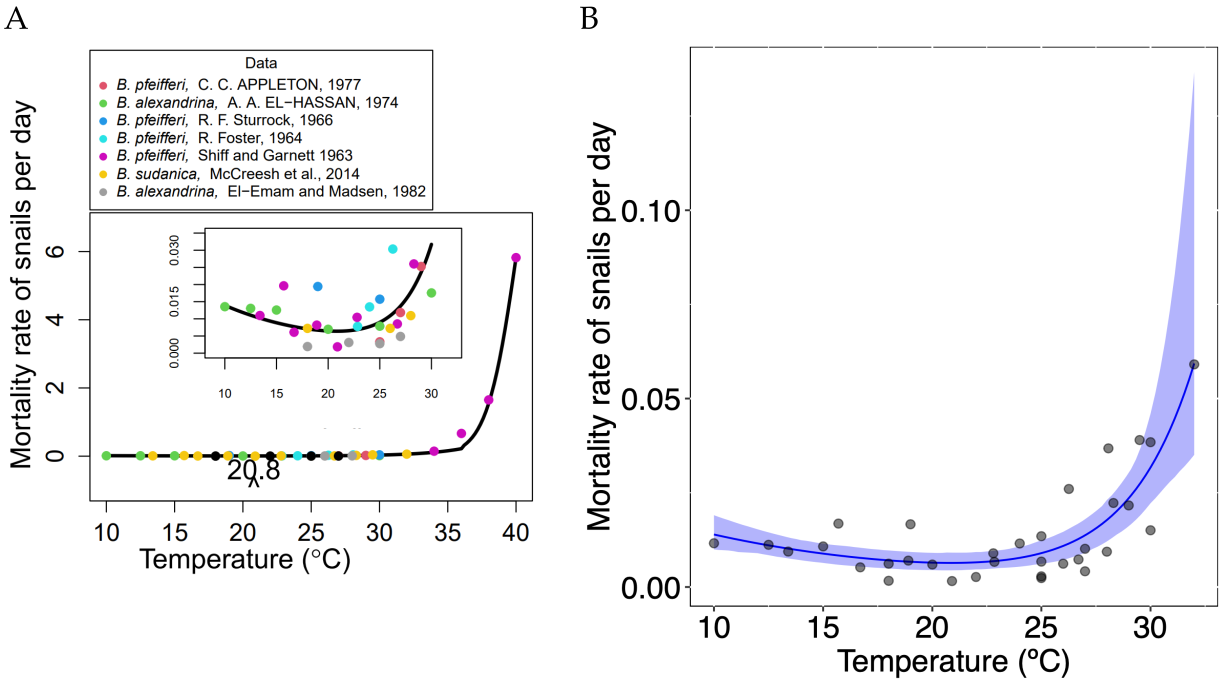

Figure 3: Mortality rate of snails and confidence interval. The different colors represent different data resources.

The top five models for *Bulinus* snails

| Model | AIC value | BIC |
|---|---|---|
| Irf | -165 | -160 |
| Modifiedgaussian | -165 | -160 |
| Joehnk | -158 | -158 |
| Spain | -158 | -153 |
| Pawar | -158 | -153 |
| Quadratic (8th in ranking) | -156 | -152 |

Quadratic curve is selected and the expression is

$$a + bT + cT^2$$

See Montagnes et al. [27] for additional information. The fitted curve to *Bulinus* data as follow

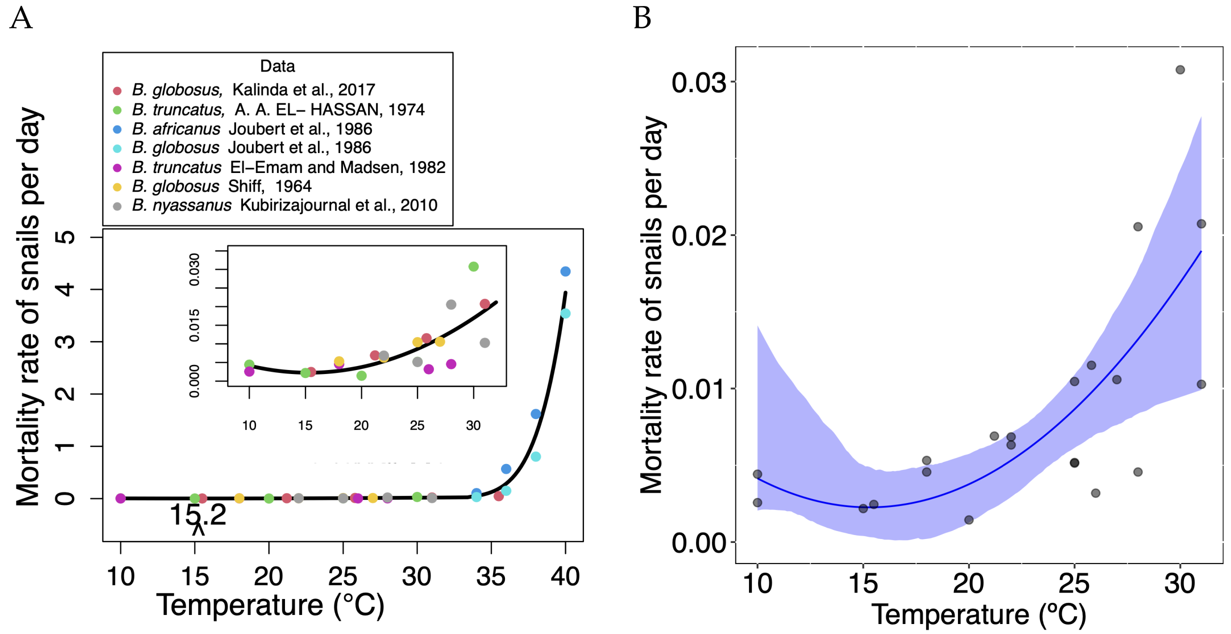

Figure 4: Mortality rate of snails and confidence interval. The different colors represent different data resources.

#### The prepatent period in snails

$\sigma_s$ has the unit of 1/day and represents the rate of snails moving from exposed class to infectious class per day.

Pfluger [28], *Biomphalaria glabrata* with *S. mansoni*. Diurnally changing water temperature.

| Temperature | Prepatent period (days) | Average of the prepatent period (days) |
|---|---|---|
| 12 hours 28-19°C, mean 24.3°C | 25-27 | 26 |
| 6 hours 28-19°C, mean 21.9°C | 35-36 | 35.5 |

| | | |
|---|---|---|
| 2 hours 28-19° C, mean 20.2° C | 43-45 | 44 |
| 1 hours 28-19° C, mean 19.8° C | 50-51 | 50.5 |
| 23.4 – 11.5° C, mean 17.5° C | 68 | 68 |
| 22.5 – 13.3° C, mean 17.9° C | 57-58 | 57.5 |
| 25.2 – 13.1° C, mean 19.2° C | 52 | 52 |
| 26.9 – 15.2° C, mean 21.1° C | 37 - 39 | 38 |
| 21.5 – 16.4° C, mean 18.9° C | 56-59 | 57.5 |
| 30 – 20° C, mean 25° C | 16-18 | 17 |
| 35.1 – 24.7$^0$ C, mean 29.9° C | 16-17 | 16.5 |
| 38.9 – 27.5° C, mean 33° C | 16-17 | 16.5 |

Pfluger [29], *Biomphalaria glabrata* with *S. mansoni*. Constant temperature

| Temperature | Prepatent period (days) |
|---|---|
| 17° C | 92.5 |
| 19° C | 55.5 |
| 22° C | 34 |
| 25° C | 24.8 |
| 28° C | 19.2 |
| 30° C | 16.3 |
| 31° C | 15.9 |
| 32° C | 14.9 |
| 33° C | 15.3 |
| 34° C | 15.9 |
| 35° C | 16 |

Foster [24], *Biomphalaria pfeifferi* with *S. mansoni*

| Temperature | Prepatent period (days) |
|---|---|
| 18° C | 57 |
| 21° C | 37 |
| 22.85° C | 32 |
| 24.01° C | 30 |
| 26.26° C | 23 |
| 28.07° C | 19 |
| 30.04° C | 18 |

| | |
|---|---|
| 31.75$^{o}$ C | 16 |

Gordon et al. [30], *Planorbis pfeifferi* with *S. mansoni*

| Temperature | Prepatent period (days) |
|---|---|
| 21.7$^{o}$ C | 36 |
| 22$^{o}$ C | 33 |
| 26.3$^{o}$ C | 23.5 |
| 26.6$^{o}$ C | 23 |
| 26.9$^{o}$ C | 22.2 |
| 27.3$^{0}$ C | 22 |
| 27.7$^{o}$ C | 19 |
| 31.8$^{o}$ C | 15.4 |
| 32.1$^{o}$ C | 15.8 |
| 32.7$^{o}$ C | 17.2 |
| 32.8$^{o}$ C | 14.7 |
| 35$^{o}$ C | 16.4 |

Gordon et al. [30], *Physopsis (bolinus) globosa* with *S. haematobium*

| Temperature | Prepatent period (days) |
|---|---|
| 22$^{o}$ C | 67 |
| 26.3$^{o}$ C | 38.5 |
| 26.4$^{o}$ C | 39.3 |

| | |
|---|---|
| 26.8° C | 34.8 |
| 31.9° C | 27.5 |
| 32.1° C | 23 |
| 33° C | 25.5 |
| 35.2° C | 26.3 |

Pfluger et al. [28], *Bulinus truncatus* with *S. haematobium*

| Temperature | Prepatent period (days) | Average of the prepatent period (days) |
|---|---|---|
| 17° C | No infection | |
| 33° C | No infection | |
| 18° C | Min 106-113, Max 120-129 | 117 |
| 19° C | Min 75-78, Max 137-151 | 110 |
| 20° C | Min 64-67, Max 91-95 | 79 |
| 21° C | Min 41-56, Max 69-75 | 60 |
| 22° C | Min 44-54, Max 62-69 | 57 |
| 23° C | Min 39-42, Max 70-76 | 56 |
| 25° C | Min 29-33, Max 52-65 | 44 |

| | | |
|---|---|---|
| 28° C | Min 22-24, Max 41-44 | 32 |
| 30° C | Min 17-19, Max 33-54 | 30 |
| 31° C | Min 17-19, Max 41-50 | 31 |
| 32° C | Min 17-20, Max 35-48 | 30 |

Yang et al. [31], *Oncomelania hupensis* with *S. japonicum*

| Temperature | Prepatent period (days) |
|---|---|
| 24° C | - N/A |
| 18° C | - |
| 21° C | 125 |
| 24° C | 90 |
| 27° C | 71 |
| 30° C | 62 |

The top five models *Biomphalaria* snails with with *S. mansoni*

| Model | AIC value | BIC value |
|---|---|---|
| Irf | -354 | -345 |
| Thomas1 | -354 | -345 |
| Briere2 | -354 | -345 |
| Ratkowsky | -354 | -345 |
| Lactin2 | -354 | -345 |

| | | |
|---|---|---|
| Gaussian (9th in ranking) | -353 | -346 |

Gaussian curve is selected and the fitted curve to *Biomphalaria* data as follow

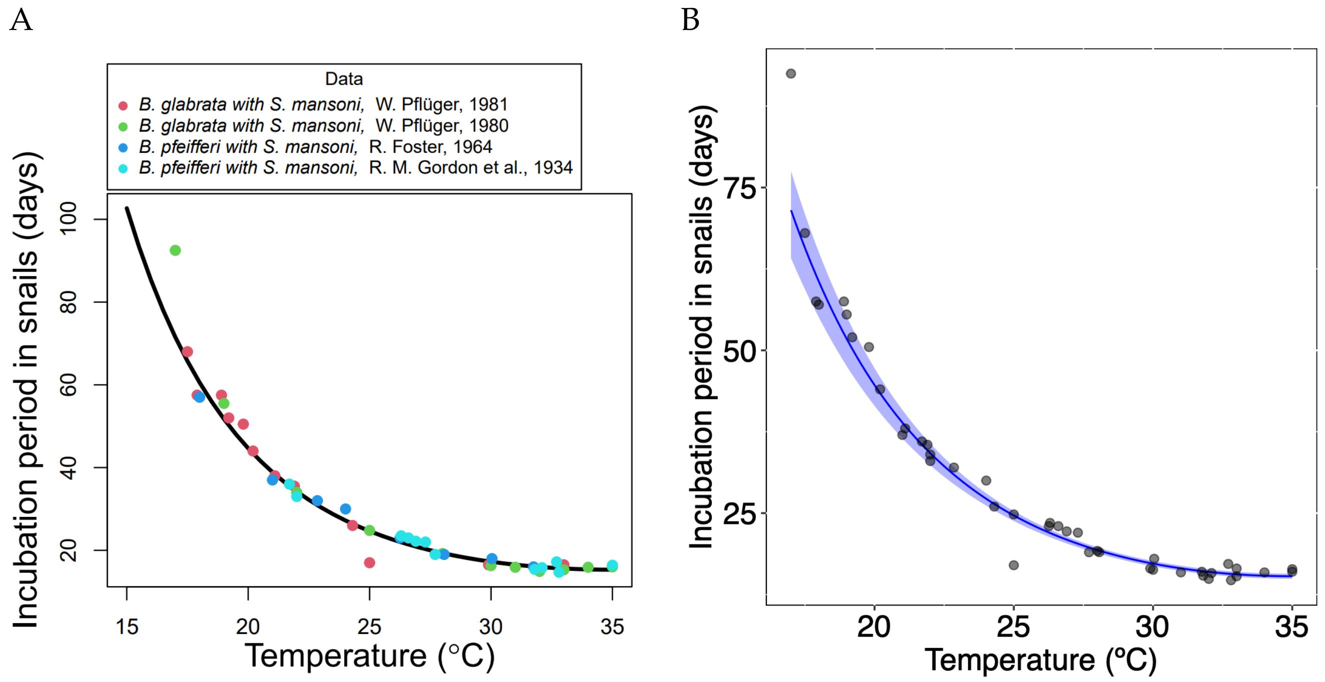

Figure 5: Prepatent period of snails and confidence interval. The different colors represent different data resources.

The top five models for *Bulinus* snails with

| Model | AIC value | BIC value |
|---|---|---|
| Gaussian | -172 | -168 |
| Lactin2 | -171 | -166 |
| Briere2 | -171 | -166 |
| Thomas1 | -170 | -166 |
| lrf | -170 | -166 |

The Gaussian curve is selected and the fitted curve to *Bulinus* data as follow

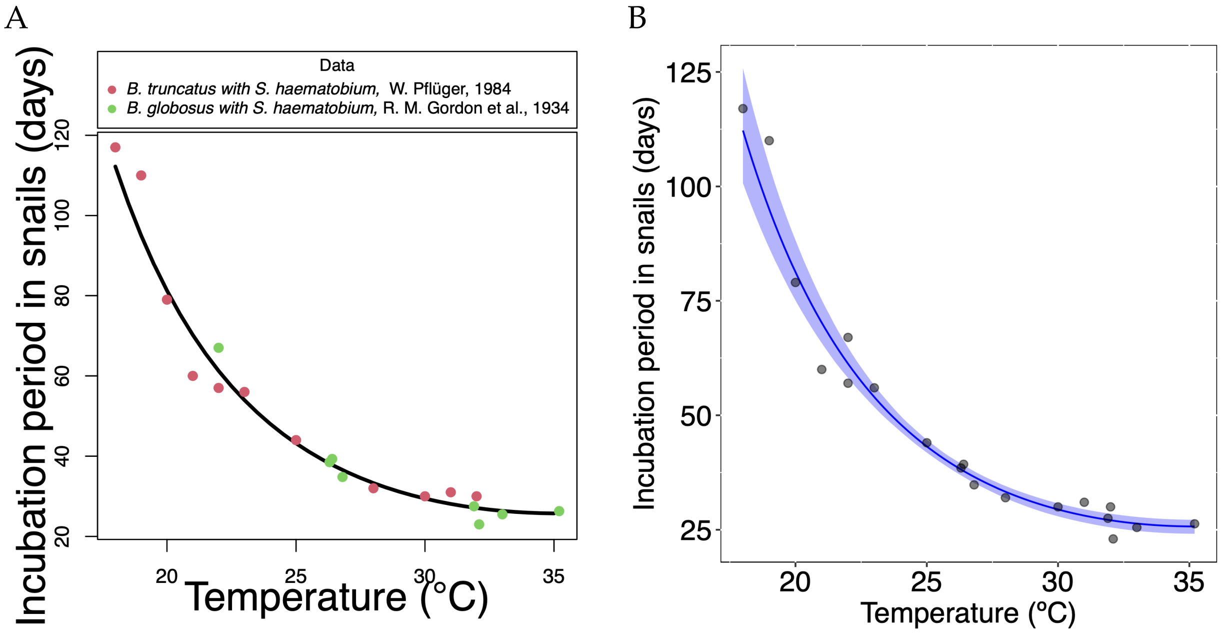

Figure 6: Prepatent period of snails and confidence interval. The different colors represent different data resources.

**The probability of hatching success of miracidia**

$\delta_e$ has the unit of parasite/egg and represents the rate of parasite hatch per egg. The only resource that we found is Nguyen et al. [32].

Nguyen et al. [33], *S. mansoni*

| Temperature | Hatching rate |
|---|---|
| 5° C | 0.2333333 |
| 9° C | 0.2 |
| 13° C | 0.3 |
| 17° C | 0.33 |
| 21° C | 0.253 |
| 22° C | 0.38 |

| | |
|---|---|
| 25$^o$ C | 0.456 |
| 29$^o$ C | 0.35 |
| 33$^o$ C | 0.5 |
| 37$^o$ C | 0.45 |

Samuelson et al. [33], *S. mansoni*

| Temperature | The half time (minutes) |
|---|---|
| $4^0$ | 120 |
| $12^0$ | 8 |
| $22^0$ | 4 |
| 34 | 2 |

The top five models

| Model | AIC value | BIC value |
|---|---|---|
| Flinn | -23.3 | -22.1 |
| Quadratic | -23 | -21.8 |
| Gaussian | -22.5 | -21.3 |
| Modifiedgaussian | -21.9 | -20.4 |
| Spain | -21.8 | -20.3 |

Flinn curve is selected and has the following expression

$$\frac{1}{1 + a + bT + cT^2}$$

See Flinn [34] for additional information. the fitted curve to *S. mansoni* data as follow

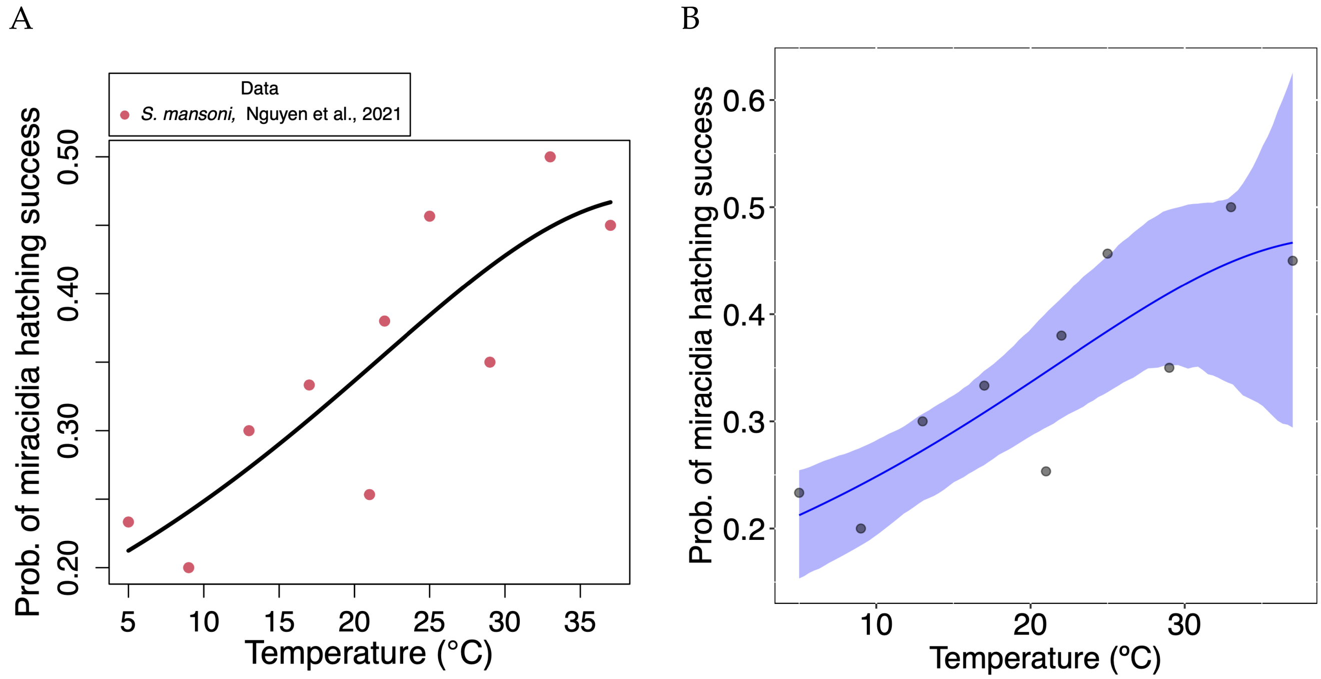

Figure 7: Probability of hatching success of miracidia and confidence interval. The different colors represent different data resources.

#### The number of cercarial released per snail

$\nu_c$ has the unit of parasite/day*snail and represents the number of cercarial produced per snail per day.

Nguyen et al. [32], *Biomphalaria glabrata* with *S. mansoni*

| Temperature | The number of cercarial releases per day per snail |
|---|---|
| 5° C | 0 |
| 9° C | 15 |
| 13° C | 260 |
| 17° C | 1350 |
| 21° C | 1970 |
| 25° C | 2900 |

| | |
|---|---|
| 29° C | 1970 |
| 33° C | 1770 |
| 37° C | 690 |

Fried et al. [35]presented data on the number of cercariae released by *Biomphalaria glabrata* during one hour under various conditions. Based on their standard (incandescent light) conditions, at different temperatures, the daily cercaria production rates (per infected snail) are:

*Biomphalaria glabrata* with *S. mansoni*

| Temperature | The number of cercarial release per hour per snail | The number of cercarial release per day per snail, adjusted based on Nguyen's results |
|---|---|---|
| 12° C | 21 | 352 $day^{-1}$ per 24 hours |
| 35° C | 350 | 5880 $day^{-1}$ per 24 hours |

Stirewalt [36], *Australorbis glabratus* with (*B. glabrata) S. mansoni.* (records are daily but from 10AM to 2PM)

| Temperature | The number of cercarial release per day per snail |
|---|---|
| 23 to 25° C | 784 |
| 26 to 28° C | 838 |

Schreiber and Schubert [37], *Australorbis glabratus (B. glabrata)* with *S. mansoni*

Temperature is 30$^o$ and the average number of cercarial produced from 9 am to 11 am is 511 when it is dark, and 692 when it is light. From 11 am to 3 pm is 340 when it is dark, 654 when it is light.

Upatham [38], *B. glabrata* with *S. mansoni*

| Temperature | Th number of cercaria after crushing the snail |
| --- | --- |
| 10$^0$ | 0 |
| 13$^0$ | 0 |
| 16$^0$ | 206 |
| 19$^0$ | 114 |
| 22$^0$ | 442 |
| 25$^0$ | 1200 |
| 28$^0$ | 1400 |
| 31$^0$ | 1233 |
| 34$^0$ | 1241 |
| 37$^0$ | 243 |
| 40$^0$ | 121 |

Kalinda et al. [18], *Bulinus globosus* with *S. haematobium*

| Temperature | The number of cercarial release per day per snail |
| --- | --- |
| 21.2$^o$ C | 1738 |
| 25.8$^o$ C | 2409 |

| | |
|---|---|
| 31° C | 829 |

The top five models for *Biomphalaria* snails

| Model | AIC value | BIC value |
|---|---|---|
| Gaussian | 314 | 318 |
| Flinn | 315 | 319 |
| Ratkowsky | 316 | 321 |
| Weibull | 316 | 321 |
| Oneill | 316 | 321 |

The Gaussian curve is selected and the fitted curve to *Biomphalaria* data as follow

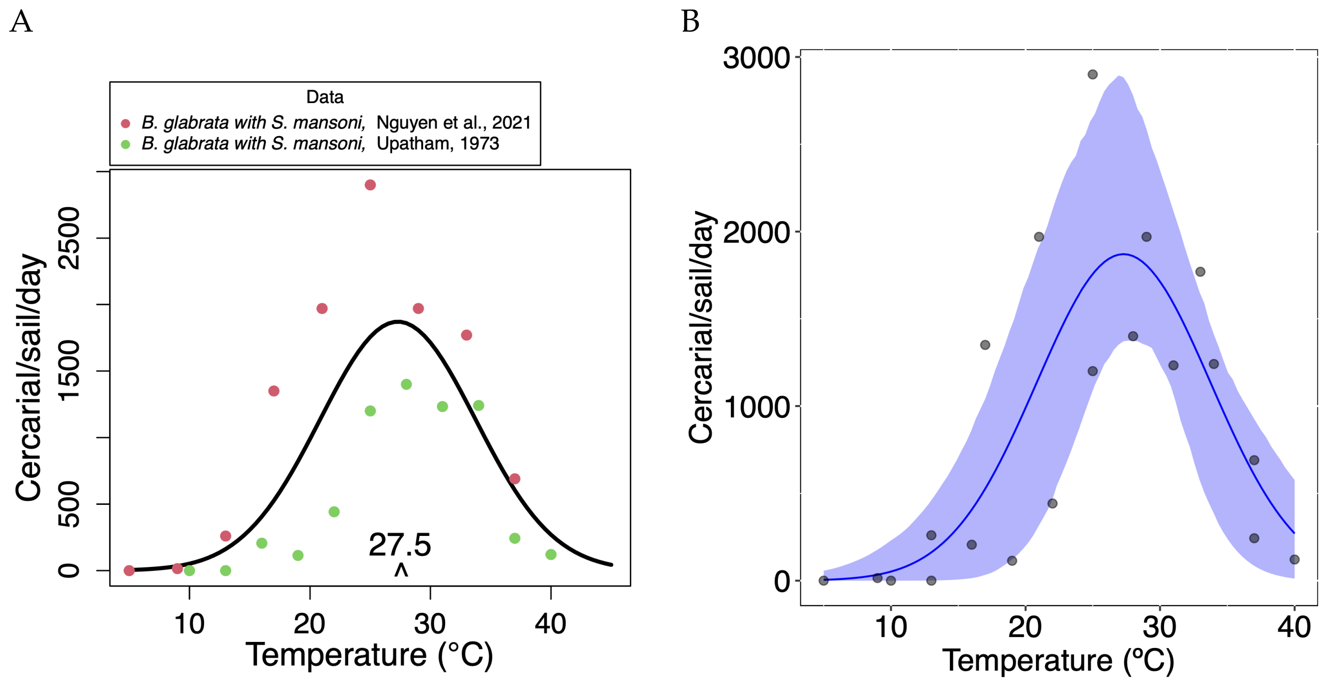

Figure 8: Cercariae release rate and confidence interval. The different colors represent different data resources.

#### The mortality rate of miracidia

$\mu_m$ has the unit of 1/day and represents the mortality rate of miracidia per day.

Anderson et al. [39], gives the life expectancy in hour for *S. mansoni*. We convert to the day unit.

| Temperature | The mortality rate of miracidia (1/day) |
|---|---|
| 5° C | 4.957653 |
| 10° C | 2.141901 |
| 15° C | 1.50839 |
| 20° C | 1.974984 |
| 25° C | 2.513615 |
| 30° C | 4.322767 |
| 35° C | 4.490178 |
| 40° C | 5.114 |

Purnell [40], experiment with *S. mansoni* and two-time range

| Temperature | 2 hours at temperature | 6 hours at temperature | The mortality rate of miracidia |
|---|---|---|---|
| 12° C | 3.261705 | 1.637893 | 2.449799 |
| 14° C | 2.396054 | 1.785148 | 2.090601 |
| 16° C | 2.904859 | 1.957561 | 2.43121 |
| 18.5° C | 1.520372 | 3.054279 | 2.287326 |
| 21.5° C | 2.337589 | 2.812790 | 2.57519 |
| 24.8° C | 2.752958 | 3.891444 | 3.322201 |

| | | | |
|---|---|---|---|
| 28.6$^o$ C | 3.842463 | 8.757026 | 6.299745 |
| 32.7$^o$ C | 4.877587 | 9.721674 | 7.299631 |

Prah and James [41] and Purnell [40] give the percentage of miracidia survival for different times at each temperature. By using these percentages and the method described in the mortality rate of snails, we found the mortality rate of miracidia as follow

Prah and James [41], *S. mansoni*

| Temperature | The mortality rate of miracidia |
|---|---|
| 5-10$^o$ C | 2.56078 |
| 18-22$^o$ C | 0.6536434 |
| 25-30$^o$ C | 1.499468 |
| 35-38$^o$ C | 3.940166 |

Prah and James [41], *S. haematobium*

| Temperature | The mortality rate of miracidia |
|---|---|
| 5-10$^o$ C | 2.275331 |
| 18-22$^o$ C | 2.210028 |
| 25-30$^o$ C | 2.24239 |
| 35-38$^o$ C | 4.354702 |

The top five models for *S. mansoni*

| Model | AIC value | BIC value |
|---|---|---|
| Thomas2 | 73.6 | 79.6 |

| | | |
|---|---|---|
| Modifiedgaussian | 74 | 78.9 |
| Spain | 74.1 | 79 |
| Quadratic | 74.2 | 78.2 |
| lrf | 75.3 | 80.3 |

Thomas2 curve is selected and has the following expression

$$ae^{bT} - (c + de^{eT})$$

see Thomas et al. [42]for more information and the fitted curve to *S. mansoni* data as follow

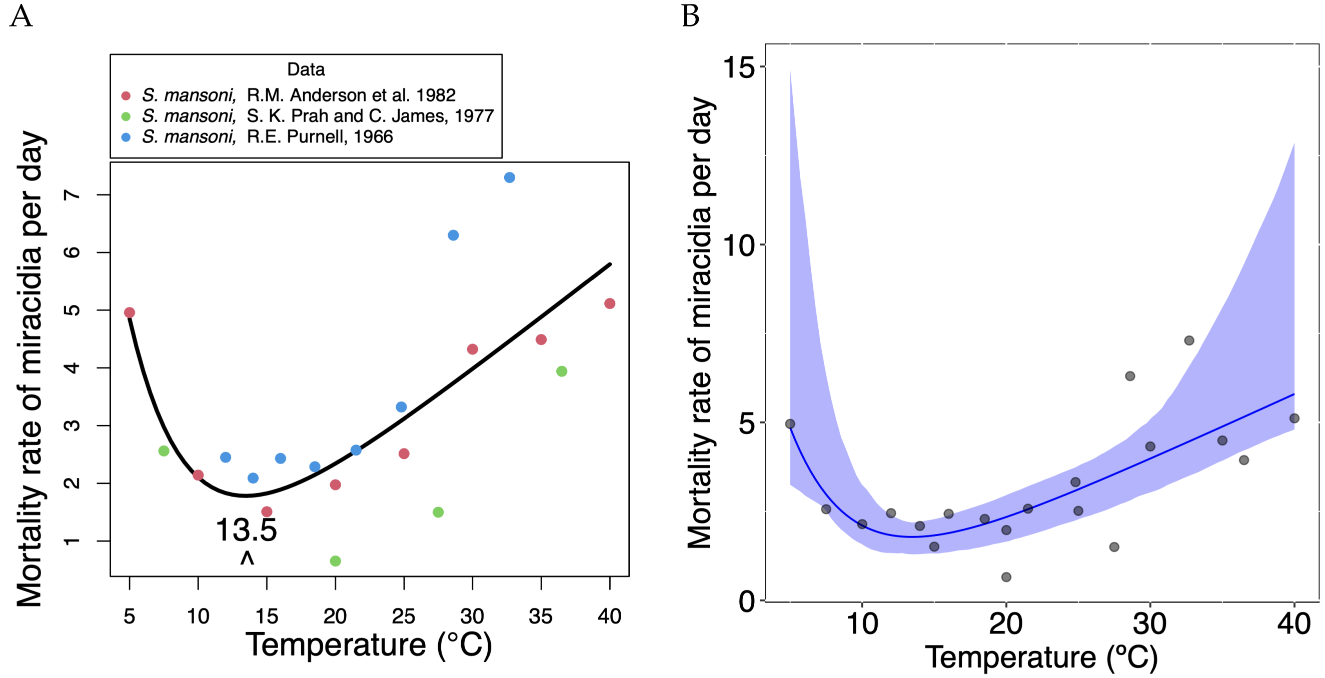

Figure 9: Mortality rate of miracidia and confidence interval. The different colors represent different data resources.

The top five models for *S. haematobium*

| Model | AIC value | BIC value |
|---|---|---|
| Joehnk | -0.509 | -0.833 |
| Spain | 0.563 | 0.292 |
| Thomas2 | 2.00 | 1.67 |

| | | |
|---|---|---|
| Quadratic | 5.57 | 5.35 |
| Modifiedgaussian | 8.62 | 8.35 |

Spain curve is selected and the fitted curve to *S. haematobium* data as follow

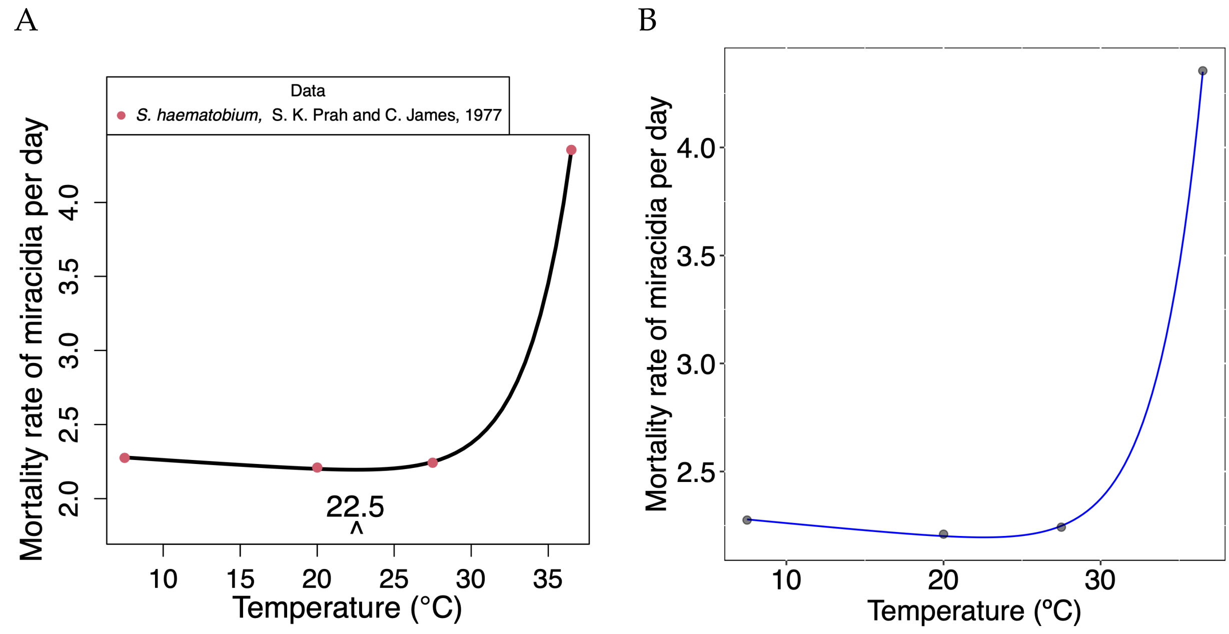

Figure 10: Mortality rate of miracidia and confidence interval. The different colors represent different data resources.

#### The mortality rate of cercaria

$\mu_c$ has the unit 1/day and represents the death rate of cercaria per day.

Lawson and Wilson [43] and Purnell [40] give the percentage of cercaria survival at different times at each temperature. By using these percentage and the method described in the mortality rate of snails, we found the mortality rate of cercaria as follow

Lawson and Wilson [43], *S. mansoni*

| Temperature | The mortality rate of cercarial |
|---|---|

| | |
|---|---|
| 10° C | 0.6844903 |
| 15° C | 0.347094 |
| 20° C | 0.9361688 |
| 25° C | 0.9384906 |
| 30° C | 1.767248 |
| 35° C | 3.79802 |
| 40° C | 7.210983 |

Purnell [40], *S. mansoni*

| Temperature | 2 hours | 6 hours | 24 hours | The mortality rate of cercarial |
|---|---|---|---|---|
| 12° C | 1.520372 | 0.9277282 | 0.5395681 | 0.9958894 |
| 15° C | 3.468196 | 2.036641 | 0.9467499 | 2.150529 |
| 18° C | 2.677723 | 1.735458 | 0.7765288 | 1.729903 |
| 21° C | 1.171354 | 1.358709 | 0.549913 | 1.026659 |
| 24° C | 1.264326 | 1.199019 | 0.7918632 | 1.085069 |
| 27° C | 1.992655 | 0.9327755 | 0.7657179 | 1.230383 |
| 30° C | 1.893889 | 1.369961 | 1.890475 | 1.718108 |
| 33° C | 1.291023 | 3.745974 | NA | NA |

Ghandour [44], *S. mansoni* (He analyze only death rate of cercaria during the penetration. We do not use in our simulaiton)

| Temperature | Mortality rate |
|---|---|

| | |
|---|---|
| $10^0$ | 9.66236 |
| $40^0$ | 10.58267 |
| 25-$27^0$ | 4.435385 |

Ghandour [44], , *S. haematobium*

| Temperature | Mortality rate |
|---|---|
| $10^0$ | 4.280099 |
| $40^0$ | 5.169395 |
| 25-$27^0$ | 1.950227 |

The top five models for *S. mansoni*

| Model | AIC value | BIC value |
|---|---|---|
| Rezende | 24.5 | 27.7 |
| Spain | 26.8 | 30 |
| Joehnk | 26.9 | 30.7 |
| Flinn | 28.2 | 30.7 |
| Thomas2 | 28.3 | 30.2 |

Spain curve is selected and the fitted curve to *S. mansoni* data as follow

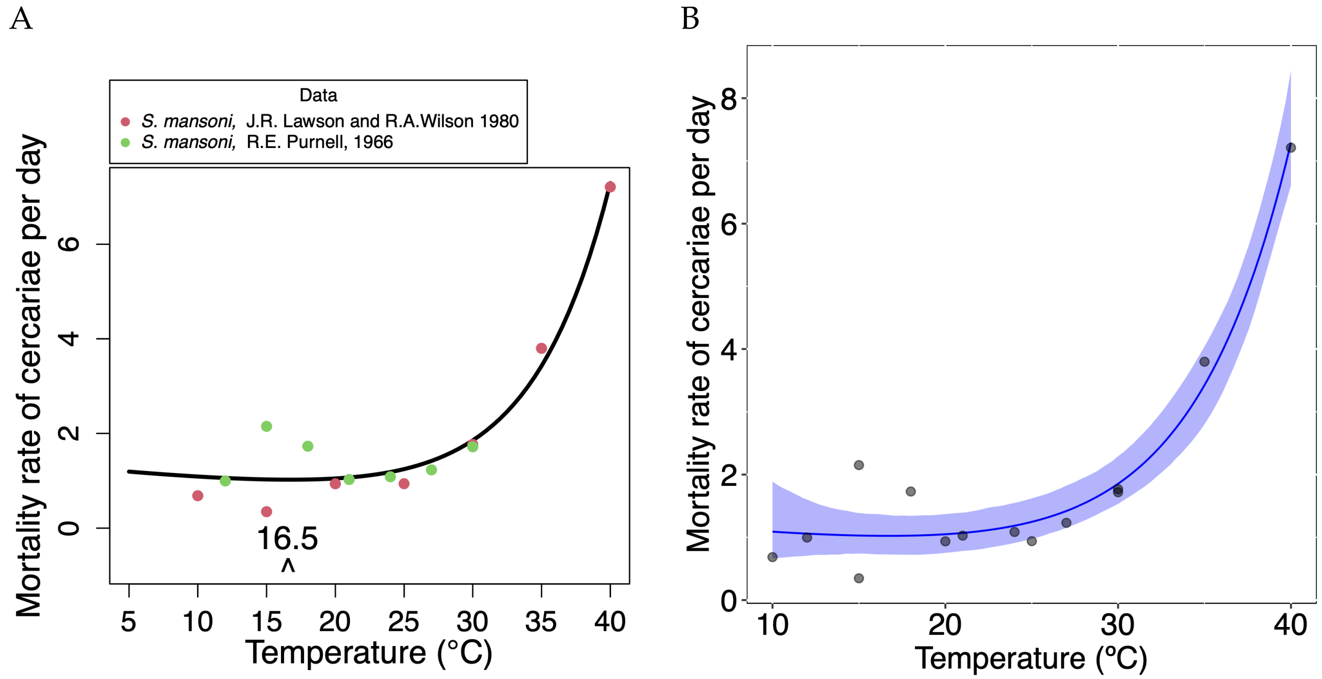

Figure 11: Mortality rate of cercaria and confidence interval. The different colors represent different data resources.

Since we do not have data for *S. haematobium*. We do not have a curve.

#### The mortality rate of infected snails

$\mu_i$ has the unit of 1/day. The value represents the death rate of snails due to infection per day.

The data is given as the percentage of death in snails after expiosed to miracidia for each day. So we use the method that we use in the mortality rate of snails to find the mortality rate of infected snails.

Pfluger [28], *Biomphalaria glabrata* and *S. mansoni*. This result from prepatent snails. This study has two experiments; the first is diurnally changing water temperature, and the second one is a sinusoidal variation of diurnal temperature.

| Temperature | Prepatent period | Average percent of mortality | Death rate |
|---|---|---|---|

| | | | |
|---|---|---|---|
| 24.3°C | (25+27)/2 = 26 (days) | (46+49)/2 = 47.5 | 0.02478296 |
| 21.9°C | 36 (days) | (43+65)/2 = 54 | 0.02157024 |
| 20.2°C | 44 (days) | (75+68)/2 = 71.5 | 0.02852877 |
| 19.7°C | 48 (days) | (66+73)/2 = 69.5 | 0.02473841 |

| Temperature | Prepatent period | Average percent of mortality | Death rate |
|---|---|---|---|
| 17.5°C | 68 (days) | 30 | 0.00524522 |
| 17.9°C | 57 (days) | 65 | 0.01841793 |
| 19.2°C | 52 (days) | 40 | 0.00982357 |
| 21.1°C | 38 (days) | 27 | 0.008281862 |
| 18.9°C | 57 (days) | (75+77)/2 = 76 | 0.02503713 |
| 25°C | 24 (days) | 28 | 0.01368767 |
| 29.9°C | 17 (days) | 30 | 0.02098088 |
| 33.1°C | 17 (days) | (18+79)/2 = 48.5 | 0.03903461 |

Pfluger [28], *Biomphalaria glabrata* and *S. mansoni*. This result from prepatent snails.

| Temperature | Prepatent perioed | Average percent of mortality | Death rate |
|---|---|---|---|

| | | | |
|---|---|---|---|
| 16$^{o}$C | 140 (days) | (95+96+98)/3 = 96.3 | 0.02354884 |
| 17$^{o}$C | 92.5 (days) | (90+58)/2 = 74 | 0.01456296 |
| 18$^{o}$C | 70 (days) | (85+65)/2 = 75 | 0.01980421 |
| 19$^{o}$C | 55.5 (days) | (57+38)/2 = 47.5 | 0.01161004 |
| 22$^{o}$C | 34 (days) | (51+64+38)/3 = 51 | 0.02098088 |
| 25$^{o}$C | 24.8 (days) | (42+58+48+22+33+20)/6 = 37 | 0.01863046 |
| 28$^{o}$C | 19.2 (days) | (9+17+33)/3 = 19.6 | 0.01136229 |
| 30$^{o}$C | 16.3 (days) | 19 | 0.01292767 |
| 31$^{o}$C | 15.9 (days) | (12+13+27)/3 = 17.3 | 0.01194658 |
| 32$^{o}$C | 14.9 (days) | (29+7+3)/3 = 13 | 0.009346447 |
| 33$^{o}$C | 15.3 (days) | (43+8)/2 = 25.5 | 0.01923994 |
| 34$^{o}$C | 15.9 (days) | (62+79)/2 = 70.5 | 0.07677861 |
| 35$^{o}$C | 16 (days) | (47+50)/2 = 48.5 | 0.04147427 |

Foster [24], *Biomphalaria pfeifferi* and *S. mansoni*

| Temperature | Death rate |
|---|---|

| | |
|---|---|
| 22.85°C | 0.006674626 |
| 24.01°C | 0.01155245 |
| 26.26°C | 0.0260108 |
| 28.07°C | 0.03678792 |

Upatham [38], *B. glabrata* with *S. mansoni*

| Temperature | Percent of survival after 12 days | Death rate |
|---|---|---|
| $10^0$ | 92.7 | 0.0058551278 |
| $13^0$ | 89.3 | 0.0085350331 |
| $16^0$ | 93 | 0.0061192732 |
| $19^0$ | 78.2 | 0.0195844770 |
| $22^0$ | 89.7 | 0.0085350331 |
| $25^0$ | 95.35 | 0.0032587267 |
| $28^0$ | 99.5 | 0.0004931228 |
| $31^0$ | 92.8 | 0.0061192732 |
| $34^0$ | 92.8 | 0.0058551278 |
| $37^0$ | 30.2 | 0.0968812480 |
| $40^0$ | 33.3 | 0.0915518574 |

Kalinda et al.[18], *Bulinus globosus* and *S. haematobium*. This result from prepatent snails.

| Temperature | Death rate |
|---|---|
| 15.5°C | 0.00244925 |
| 21.2°C | 0.006901087 |
| 25.8°C | 0.01151302 |

| | |
|---|---|
| 31°C | 0.02073716 |
| 36°C | 0.04642702 |

Pfluger et al. [29], *Bulinus truncatus* and *S. haematobium*. This result from prepatent snails.

| Temperature | Length of prepatent | Average | Percentage of death | Death rate |
|---|---|---|---|---|
| 17°C | No infection | - | - | - |
| 33°C | No infection | - | - | - |
| 18°C | Min 106-113, Max 120-129 (days) | [109.5, 124.5] = 117 | 78.6 | 0.0131776 |
| 19°C | Min 75-78, Max 137-151 (days) | [76.5, 144] = 110.25 | 71.6 | 0.01141752 |
| 20°C | Min 64-67, Max 91-95 (days) | [65.5, 93] = 79.25 | 78.9 | 0.01963277 |
| 21°C | Min 41-56, Max 69-75 (days) | [48.5, 72] = 60.25 | 72.3 | 0.02130685 |
| 22°C | Min 44-54, Max 62-69 (days) | [49, 65.5] = 57.25 | 71.7 | 0.02204905 |
| 23°C | Min 39-42, Max 70-76 (days) | [40.5, 73] = 56.75 | 77.6 | 0.02636316 |
| 25°C | Min 29-33, Max 52-65 (days) | [31, 58.5] = 44.75 | 62.4 | 0.02185846 |

| | | | | |
|---|---|---|---|---|
| 28°C | Min 22-24, Max 41-44 (days) | [23, 42.5] = 32.75 | 40.6 | 0.01590461 |
| 30°C | Min 17-19, Max 33-54 (days) | [18, 43.5] = 30.75 | 38.3 | 0.01570362 |
| 31°C | Min 17-19, Max 41-50 (days) | [18, 45.5] = 31.75 | 54.4 | 0.02473268 |
| 32°C | Min 17-20, Max 35-48 (days) | [18.5, 41.5] = 30 | 54.5 | 0.0262486 |

Chu et al. [45] *Bulinus truncatus* with *S. haematobium*

| Temperature | Number of exposed snails | Number of snails alive on the 50th day after exposure | Death rate |
|---|---|---|---|
| $10^0$ | 100 | 85 | 0.003250379 |
| $12^0$ | 50 | 40 | 0.004462871 |
| $14^0$ | 50 | 42 | 0.003487068 |
| $15^0$ | 100 | 90 | 0.00210721 |
| $20^0$ | 100 | 80 | 0.004462871 |
| $25^0$ | 150 | 118 | 0.004799013 |
| $30^0$ | 97 | 87 | 0.002176057 |
| $35^0$ | 97 | 89 | 0.001721492 |
| $38^0$ | 45 | 37 | 0.003914892 |

The top five models for *Biomphalaria snails*

| Model | AIC value | BIC value |
|---|---|---|
| Joehnk (1st) | -223 | -213 |
| Spain (2nd) | -219 | -211 |
| Thomas2 (3rd) | -218 | -208 |
| Irf (4th) | -216 | -208 |
| Modifiedgaussian (5th) | -213 | -205 |

Flinn curve is selected and  the fitted curve to *Biomphalaria* data as follow

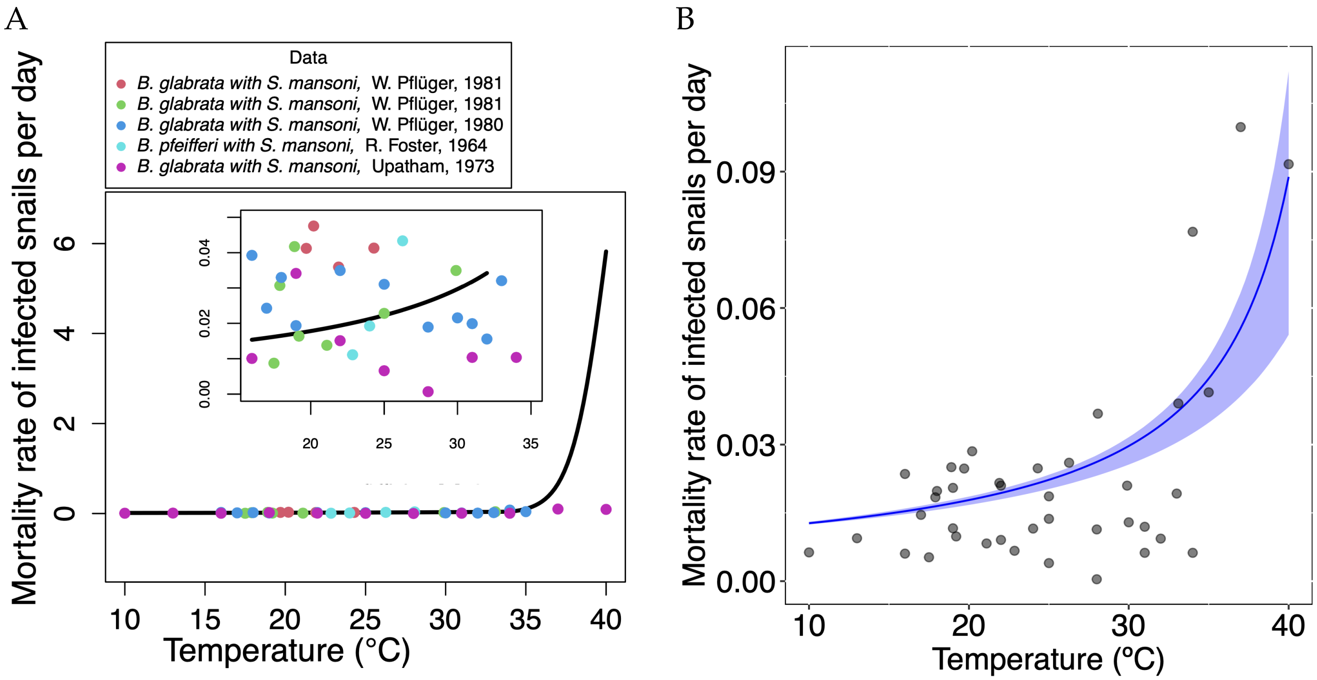

Figure 12: Mortality infected snails and confidence interval. The  different colors represent different data resources.

The top five models for *Bulinus* snails.

| Model | AIC value | BIC value |
|---|---|---|
| Joehnk | -173 | -165 |

| | | |
|---|---|---|
| Weibull | -160 | -154 |
| Spain | -152 | -146 |
| Thomas | -151 | -143 |
| Irf | -149 | -143 |

Spain curve is selected and the fitted curve to *Bulinus* data as follow

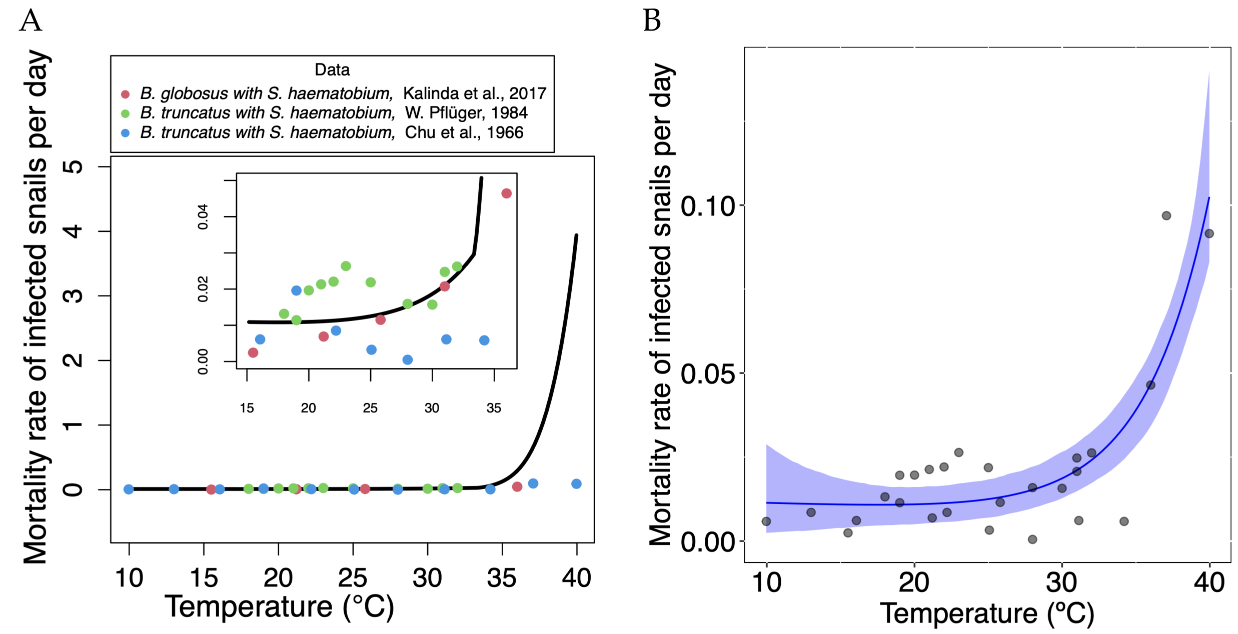

Figure 13: Mortality infected snails and confidence interval. The different colors represent different data resources.

#### The transmission rate of schistosomiasis in snails

$\beta_s$ has the unit of snail/parasite and represents the number of parasites per snail

$\beta_s$ varies with many environmental factors, such as density of miracidia per a snail or habitat for snails, so it is hard to find a good estimate of $\beta_s$. However, since the main focus of this study is the thermal curve, we can approximate the shape of this thermal curve by using the percentage of infected snails from different experiments. To do that, we use a simpler model used in (Mangal et al. [46])

$$\frac{dU}{dt} = -MU\beta_s$$

$$\frac{dI}{dt} = MU\beta_s$$

$$\frac{dM}{dt} = -MN\beta_s - \mu_m M$$

where $U$ are the number of uninfected snails, $I$ the number of infected snails, $N$ the total number of snails $(U+I)$ and $M$ the number of miracidia. After we solve this dynamic we get $\beta_s$

Given $M_0$ is the initial number of miracidia, $t$ is the day.

Stirewalt [36], *Australorbis glabratus* with *S. mansoni.* A group of twenty to twenty-five snails exposed one miracidium each at time for 24 hours individually.

| Temperature | Infection rate (%) | $\beta_s$ value |
|---|---|---|
| 23-25°C | 32 | 0.07515584 |
| 26-28°C | 54 | 0.4808139 |
| 31-33°C | 50 | 0.3922359 |

Foster [24], *Biomphalaria pfeifferi* with *S. mansoni.* Snail exposed four miracidia and 30 snails were used in the each experiment for 24 hours.

| Temperature | Infection rate (%) | $\beta_s$ value |
|---|---|---|
| 22.85°C | 21/30 = (70) | 0.04079299 |
| 24.01°C | 21/30 = (70) | 0.04333207 |
| 26.26°C | 28/30 = (93) | 0.2203394 |
| 28.07°C | 24/30 = (80) | 0.08210066 |
| 30.04°C | 21/30 = (70) | 0.05756868 |
| 31.75°C | 23/30 = (76) | 0.07958718 |

Prah and James [41], *Biomphalaria pfeifferi* with *S. mansoni.* Four to eight snails exposed ten miracia for 3 to 6 hours.

| Temperature | Infection rate (%) | $\beta_s$ value |
|---|---|---|
| 5º C | 0.5 | 0.0006344901 |
| 15º C | 40 | 0.05059652 |
| 20º C | 77 | 0.1629059 |
| 22.5º C | 85 | 0.2249733 |

Anderson et al. [39], *Biomphalaria glabrata* with *S. mansoni.* Single snail exposed five miracidia for one hour replication 30 snails.

| Temperature | Infection rate (%) | $\beta_s$ value |
|---|---|---|
| 15º C | 83 | 0.3658224 |
| 20º C | 84 | 0.3865833 |
| 25º C | 96 | 0.9074138 |
| 30º C | 88 | 0.487644 |
| 35º C | 84 | 0.4107836 |

DeWitt [47], *B. glabrata* with *S. mansoni.* Each snail exposed to five miracidia for 2 hours.

| Temperature | Dominican Republic; Surv./%Pos | BSurv./Pos razil; Surv./%Pos | Puerto Rico Surv./%Pos | Venezuela (1946) Surv./%Pos | Venezuela (1952) Surv./%Pos |
|---|---|---|---|---|---|
| $10^0$ | 62/0 (0) | - | 88/0 (0) | 86/0 (0) | 75/0 (0) |
| $25^0$ | 142/46 (0.0078523) | 48/0 (0) | 76/43 (0.032993) | 82/27 (0.01388562) | 82/68 (0.07405795) |
| $35^0$ | 116/73 (0.0283053) | 52/0 (0) | 100/53 (0.0241807) | 67/36 (0.03692433) | 68/80 (0.09039597) |

| | | | | | |
|---|---|---|---|---|---|
| $40^0$ | 5/0 (0) | 12/0 (0) | 0/0 (0) | 12/58 (0.01246124) | 10/90 (0.00401955) |

Coelho and Bezerra [48], *Biomphalari glabrata* with *S. mansoni*. 30 snails individually exposed to 10 miracidia for 12 hours.

| Temperature | The percent of snails positive | $\beta_s$ value |
|---|---|---|
| $15^0$ | 1.3 | 0.0001342012 |
| $20^0$ | 46 | 0.007285127 |
| $30^0$ | 89 | 0.04153248 |

Upatham et al. [38], *B. glabrata* with *S. mansoni.* 360 snails exposed to four miracidia indivudully for 1 hours.

| Temperature | Percent of positive | $\beta_s$ value |
|---|---|---|
| $10^0$ | 0 | 0 |
| $13^0$ | 0 | 0 |
| $16^0$ | 15 | 0.002895729 |
| $19^0$ | 21.5 | 0.004360468 |
| $22^0$ | 40.5 | 0.009807099 |
| $25^0$ | 48.6 | 0.01299096 |
| $28^0$ | 50 | 0.01373957 |
| $31^0$ | 53 | 0.01528639 |
| $34^0$ | 70.65 | 0.02722317 |
| $37^0$ | 13.43 | 0.002743025 |

| | | |
|---|---|---|
| $40^0$ | 9.95 | 0.001999862 |

Prah and James [41], *Bulinus (Physopsis) globosus* with *S. haematobium.* Four to eight snails exposed ten miracidia for 3 to 6 hours.

| Temperature | Infection rate (%) | $\beta_s$ value |
|---|---|---|
| $5^o$ C | 1 | 0.001279126 |
| $12^o$ C | 37 | 0.04565688 |
| $19^o$ C | 64 | 0.1084776 |
| $26^o$ C | 78 | 0.1849867 |

Shiff [49], *Bulinus globosus* infected with *S. haematobium.* 10 snails exposed 200 miracidia for each replicated for 24 hours in each experiment

| Temperature | Percent of infection | $\beta_s$ value |
|---|---|---|
| $25^0$ | 87.4 | 0.03730699 |
| $20^0$ | 23.5 | 0.003512327 |
| $18^0$ | 56.4 | 0.01022082 |
| $16^0$ | 14.8 | 0.001782906 |
| $13^0$ | 21 | 0.002553961 |
| $11^0$ | 10.5 | 0.001281865 |

Chu et al. [45], *Bulinus trancatus* infected with *S. haematobium.* Two miracidia were used in each tube for two hours.

| Temperature | Number of exposed snails | Total number of snails found positive after the 50th day | $\beta_s$ value |
|---|---|---|---|
| $10^0$ | 100 | 3 | 0.002009499 |
| $12^0$ | 50 | 3 | 0.008145561 |
| $14^0$ | 50 | 7 | 0.02029471 |
| $15^0$ | 100 | 25 | 0.02016721 |
| $20^0$ | 100 | 45 | 0.04754069 |
| $25^0$ | 150 | 71 | 0.03577785 |
| $30^0$ | 97 | 49 | 0.06475144 |
| $35^0$ | 97 | 38 | 0.04395093 |
| $38^0$ | 45 | 12 | 0.05660857 |

The top five models for *Biomphalaria* snails with *S. mansoni*

| Model | AIC value | BIC value |
|---|---|---|
| Sharpeschoollow | 13.7 | 26.8 |
| Pawar | 14.7 | 24 |
| Sharpeschoolfull | 14.7 | 24 |
| Ratkowsky | 14.7 | 24.1 |
| Briere2 | 15.1 | 24.4 |
| Spain (12th in ranking) | 17 | 26.4 |

Spain curve is selected and the fitted curve to *Biomphalaria* data as follow

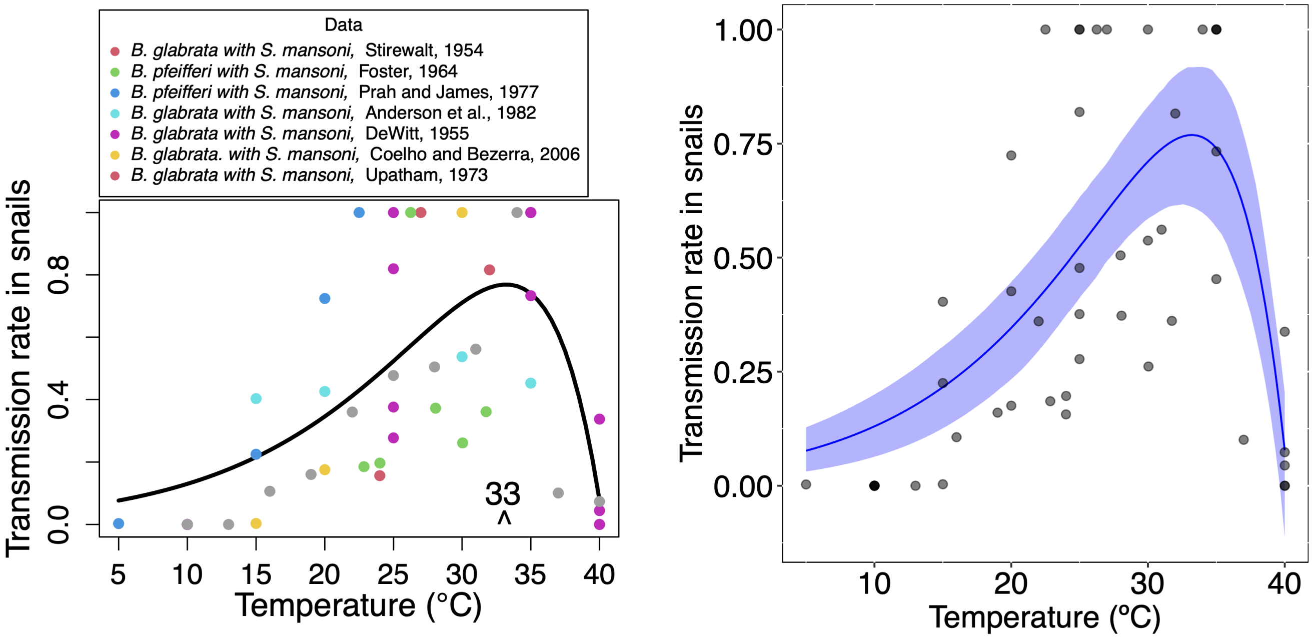

Figure 14: Transmission rate in snails and confidence interval. The different colors represent different data resources.

The top five models for *Bulinus trancatus* with *S. haematobium*

| Model | AIC value | BIC value |
|---|---|---|
| Ratkowsky | -5.84 | -1.12 |
| Gaussian | -5.75 | -1.97 |
| Sharpeschoo lhigh | -4.54 | 0.181 |
| Pawar | -4.54 | 0.181 |
| Weibull | -3.87 | 0.850 |
| Flin (8th in ranking) | -3.23 | 0.548 |

Flinn curve is selected and the fitted curve to *Bulinus* data as follow

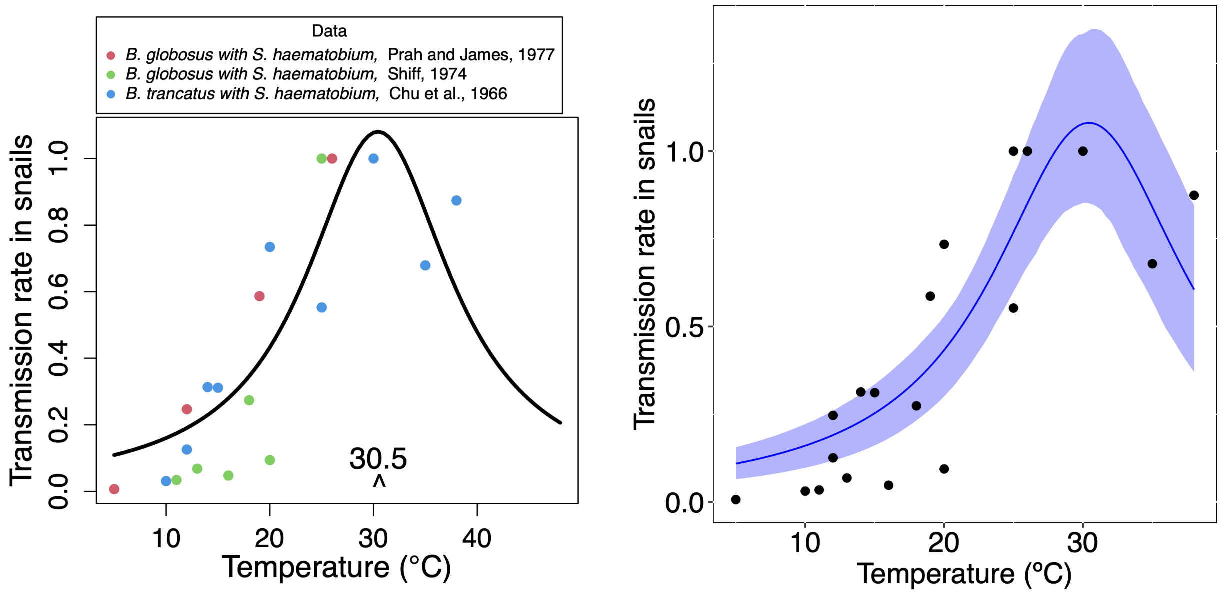

Figure 15: Transmission rate in snails and confidence interval. The different colors represent different data resources.

#### The transmission rate of schistosomiasis in humans

$\beta_h$ has the unit of 1/human*day and represents the rate of parasites that penetrate human skin per day. We use a model used in Mangal et al. [46]to adjust $\beta_h$

$$\frac{dC(t)}{dt} = -\beta_h C(t)M - \mu_c C(t)$$

$$\frac{dW(t)}{dt} = \beta_h C(t)M$$

where $C$ is the number of cercariae in the water and $M$ is the number of mice (which remained constant). Thus if we solve for $\beta_h$ we can find the transmission rate of disease in snails.

Purnell [40], *S. mansoni,* Six hamsters expose to 200 original cercariae for one hour.

| Temperature | 2 hours percent | 6 hours percent | 24 hours percent |
|---|---|---|---|

| | penetrate rate ($\beta_h$) | penetrate rate ($\beta_h$) | penetrate ($\beta_h$) |
|---|---|---|---|
| 12° C | 18.5 (0.9246078) | 17.4 (1.0535) | 14.3 (2.182979) |
| 15° C | 16.5 (0.810981) | 16.8 (1.002849) | 11.1 (1.523512) |
| 18° C | 25.3 (1.320736) | 15.7 (0.9284408) | 12.6 (1.777458) |
| 21° C | 21.4 (1.092269) | 23.2 (1.481275) | 12.4 (1.840471) |
| 24° C | 14.2 (0.6998873) | 12 (0.7225294) | 5.6 (0.8305635) |
| 27° C | 22.9 (1.231836) | 10.3 (0.6554545 | 3.8 (0.7054314) |
| 30° C | 14.8 (0.7901292) | 16.3 (1.251547) | 4.3 (1.339291) |
| 33° C | 11.1 (0.6292689) | 8.4 (0.7475422) | 0 (0) |

Stirewalt [36], *S. mansoni.* Each mouse was exposed to 50 cercariae and repeated for 100 mice. We average the number of cercariae that successfully penetrate mice's skin for one hour.

| Temperature (°C) | The number of cercaria convert to worm | Cercarial transmission rate ($\beta_h$) |
|---|---|---|
| 23-25 | 1610 | 0.09590499 |
| 26-28 | 2320 | 0.1552625 |

DeWitt [47] with *S. mansoni*

Each mouse exposed to 67 cercaria for 30 minutes and repeated with15 mices.

| Temperature | Percentage of cercarial infected | Cercarial transmission rate ($\beta_h$) |
|---|---|---|
| $10^0$ | ND | |
| $15^0$ | 13.3 | 0.4618475 |
| $20^0$ | 23 | 0.8464159 |
| $25^0$ | 26.7 | 1.00836 |
| $30^0$ | 31.5 | 1.237522 |
| $35^0$ | 17 | 0.6192221 |
| $40^0$ | ND | |
| $45^0$ | ND | |

12 mice and average cercaria per mouse 47 for 30 munites.

| Temperature | Percentage of cercarial infect | Cercarial transmission rate ($\beta_h$) |
|---|---|---|
| $10^0$ | ND | |
| $15^0$ | 9.1 | 0.385854 |
| $20^0$ | 15.7 | 0.6910937 |
| $25^0$ | 14.5 | 0.635236 |
| $30^0$ | 21.3 | 0.9783329 |

| | | |
|---|---|---|
| $35^0$ | 22.6 | 1.0652 |
| $40^0$ | 6.2 | 0.2764123 |
| $45^0$ | ND | |

10 mice exposed to 70 cercaria in average for 30 munites.

| Temperature | Percentage of cercarial infect | Cercarial transmission rate ($\beta_h$) |
|---|---|---|
| $10^0$ | 14.2 | 0.7439515 |
| $15^0$ | 30.4 | 1.76077 |
| $20^0$ | 40 | 2.484149 |
| $25^0$ | 51.2 | 3.50177 |
| $30^0$ | 55.3 | 3.9655 |
| $35^0$ | 49.4 | 3.421058 |
| $40^0$ | 13.5 | 0.7532138 |
| $45^0$ | 0 | 0 |

12 mice exposed to 76 cercaria in average for 30 minutes.

| Temperature | Percentage of cercarial infect | Cercarial transmission rate ($\beta_h$) |
|---|---|---|
| $0^0$ | 0 | 0 |
| $5^0$ | 0 | 0 |
| $10^0$ | 2.9 | 0.1190698 |

| | | |
|---|---|---|
| $15^0$ | 14.8 | 0.6479505 |
| $20^0$ | 29.2 | 1.39834 |
| $25^0$ | 30.9 | 1.50036 |
| $30^0$ | NA | |
| $35^0$ | ND | |
| $40^0$ | ND | |
| $45^0$ | ND | |

The top five models for *Biomphalaria* snails with *S. mansoni*

| Model | AIC value | BIC value |
|---|---|---|
| Modifiedgaussian | 3.05 | 12.7 |
| Sharpeschoolfull | 3.83 | 17.4 |
| Quadratic | 4.38 | 12.1 |
| lrf | 5.54 | 15.2 |
| Gaussian | 5.95 | 13.7 |
| Briere (8th in ranking) | 6.86 | 16.5 |

Briere curve is selected and has the following expression

$$aT(T - T_{min})(T_{max} - T)^{\frac{1}{b}}$$

See Briere et al. [50] for additional information and the fitted curve to *Biomphalaria* data as follow

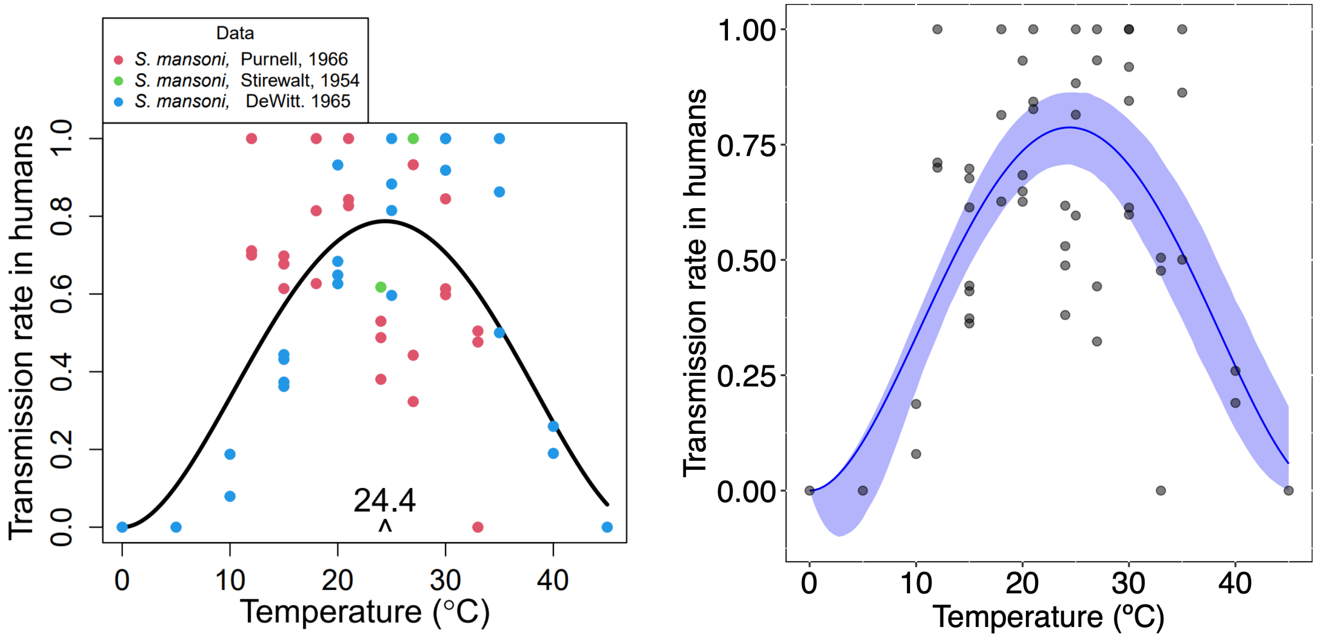

Figure 16: Transmission rate in humans and confidence interval. The different colors represent different data resources.

### Basic reproduction number

#### The calculation of basic reproduction number

We use the next generation matrix method developed by Driessche and Watmough [51] to calculate $\mathcal{R}_0$. The method gives a shortcut to draw $\mathcal{R}_0$ value. First, we find the rates of new infection for all infected compartments. We call this $\mathcal{F}$.

$$\mathcal{F} = \begin{bmatrix} \lambda(T)S \\ 0 \\ \beta_h(T)\dfrac{\nu_c(T)}{\mu_c(T)}I \\ 0 \\ 0 \end{bmatrix}$$

Then, we find the rates of changes for all other compartments. We call this $\mathcal{V}$.

$$\mathcal{V} = \begin{bmatrix} (\sigma_s(T) + \mu_i(T))E \\ -\sigma_s(T)E + \mu_i(T)I \\ \sigma_p W \\ -\sigma_p W + (\mu_h + \mu_p)W_m \\ -(\nu_s(T) - (S + E + I)\nu)(S + rE) + (\lambda(T) + \mu(T))S \end{bmatrix}$$

We calculate the Jacobian matrix from new infected compartment to all infectious compartments. We call this **F**.

$$\mathbf{F} = \begin{bmatrix} 0 & 0 & 0 & \dfrac{\Lambda\beta_s(T)h\delta_e(T)\nu_e S}{\mu_m(T)(S + E + I)} \\ 0 & 0 & 0 & 0 \\ 0 & \dfrac{\beta_h(T)\nu_c(T)}{\mu_c(T)} & 0 & 0 \\ 0 & 0 & 0 & 0 \end{bmatrix}$$

We calculate the Jacobian of matrix from all infectious compartment to all infectious compartments. We call this **V**.

$$\mathbf{V} = \begin{bmatrix} \sigma_s(T) + \mu_i(T) & 0 & 0 & 0 \\ -\sigma_s(T) & \mu_i(T) & 0 & 0 \\ 0 & 0 & \sigma_p & 0 \\ 0 & 0 & -\sigma_p & \mu_h + \mu_p \end{bmatrix}$$

We need to calculate the inverse of **V** to find the next generation matrix.

$$\mathbf{V^{-1}} = \begin{bmatrix} \dfrac{1}{\sigma_s(T) + \mu_i(T)} & 0 & 0 & 0 \\ \dfrac{\sigma_s(T)}{\mu_i(T)(\sigma_s(T) + \mu_i(T))} & \dfrac{1}{\mu_i(T)} & 0 & 0 \\ 0 & 0 & \dfrac{1}{\sigma_p} & 0 \\ 0 & 0 & \dfrac{1}{\mu_h + \mu_p} & \dfrac{1}{\mu_h + \mu_p} \end{bmatrix}$$

The product of $\mathbf{FV^{-1}}$ at DFE equilibrium gives us the next generation matrix and the spectral radios of the next generation matrix equals to a threshold value of the basic reproduction number $\mathcal{R}_0$

$\mathbf{FV^{-1}(X^*)} =$

$$\begin{bmatrix} 0 & 0 & \frac{\Lambda\beta_s(T)h\delta_e(T)\nu_e S^*}{\mu_m(T)(\mu_h+\mu_p)S^*} & \frac{\Lambda\beta_s(T)h\delta_e(T)\nu_e S^*}{\mu_m(T)(\mu_h+\mu_p)S^*} \\ 0 & 0 & 0 & 0 \\ \frac{\beta_h(T)\nu_c(T)\sigma_s(T)}{\mu_c(T)\mu_i(T)(\sigma_s(T)+\mu_i(T))} & \frac{\beta_h(T)\nu_c(T)}{\mu_c(T)\mu_i(T)} & 0 & 0 \\ 0 & 0 & 0 & 0 \end{bmatrix}$$

Note we evaluate the next generation matrix at the DFE $X^* = (S^*, 0,0,0,0)$. We can find a disease free equilibrium (DFE) by simple setting the right hand side of the Ordinary differential equation (ODE) system to zero and plugging the DFE. Therefore,

$$(\nu_s(T) - \mu(T) - S^*\nu)S^* = 0 \Rightarrow S^* = \frac{\nu_s(T) - \mu(T)}{\nu}$$

The DFE is

$$(S^*, E^*, I^*, W^*, W_m^*) = \left(\frac{\nu_s(T) - \mu(T)}{\nu}, 0,0,0,0\right)$$

Calculate the spectral radios of the next generation matrix we get the basic reproduction number $\mathcal{R}_0$. Note that we assume $\nu_s(T) - \mu(T) > 0$ in order to biologically be mean-full.

$$\mathbf{FV^{-1}(X^*)} = \mathcal{R}_0 = \left(\frac{\Lambda\beta_s(T)h\delta_e(T)\nu_e\beta_h(T)\nu_c(T)\sigma_s(T)}{\mu_m(T)(\mu_h+\mu_p)\mu_c(T)\mu_i(T)(\sigma_s(T)+\mu_i(T))}\right)^{1/2}$$

### Model comparison with GNTD data

We subset the data to only include non-zero data points to focus on areas where schistosomiasis is endemic.

#### GNTD prevalence data and temperature

We fit a quantile GAM regression using the qgam R package to explore the relationship between temperature and schistosomiasis prevalence.

#### R0 as a predictor of prevalence

We used quantile regressions to evaluate our R0 estimate as a predictor of prevalence. We found that our estimate of R0 is a significant predictor of prevalence for all quantiles considered ($\tau$ =0.25,0.5,0.75, 0.9, 0.95, 0.98, p-value < 0.005 for all, Figure 17). The R0 estimate from Nguyen et al. was not a significant predictor of prevalence, except for the 25th quantile. In addition, the model using our R0 estimate had a lower AICc (AIC(Aslan et al) = 3240, AIC(Nguyen et al) = 3297, $\tau$ = 0.5).

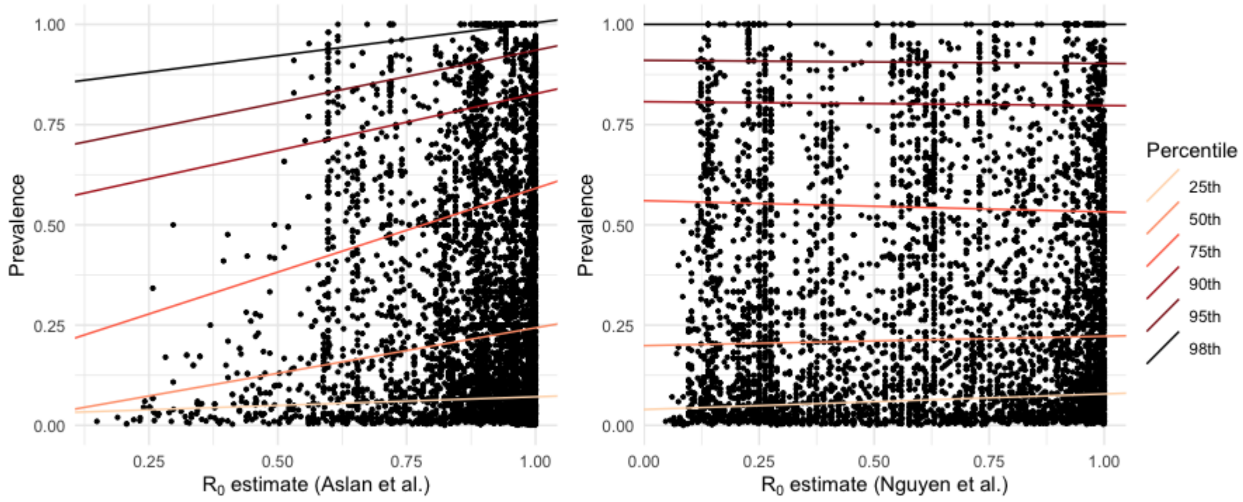

Figure 17 Quantile regression of prevalence over R0 estimates for our estimate and for Nguyen et al. estimate.

### Human water contact rate

We formulate water contact rate for varying temperature by using a sigmoid function as follow

$$C(T) = \frac{1}{1 + e^{-a(T - T_{med})}} + b$$

[52]

### Sensitivity

$$\frac{d\mathcal{R}_0}{dT} = \frac{\partial \mathcal{R}_0}{\partial x_1}\frac{\partial x_1}{\partial T} + \ldots + \frac{\partial \mathcal{R}_0}{\partial x_n}\frac{\partial x_n}{\partial T}$$

if we are interested in on certain parameters. We can simple assume all other parameter are constant, so

$$\frac{d\mathcal{R}_0}{dT} = \frac{\partial \mathcal{R}_0}{\partial x}\frac{\partial x}{\partial T}$$

### The partial derivative of each function

if we take the derivative of each thermal curve function,

- Gaussian; The explicit equation is

$$G(T) = r_{max}e^{-0.5\left(\frac{T-T_{opt}}{a}\right)^2}$$

Then the derivative would be

$$\frac{dG(T)}{dT} = \frac{-r_{max}\left(T - T_{opt}\right)}{a^2}e^{-0.5\left(\frac{T-T_{opt}}{a}\right)^2}$$

- Spain; The explicit equation is

$$S(T) = r_0 e^{aT}(1 - be^{cT})$$

Then the derivative would be

$$\frac{dS(T)}{dT} = r_0 e^{aT}(a - abe^{cT} - bce^{cT})$$

- Filnn; The explicit equation is

$$F(T) = \frac{1}{1 + a + bT + cT^2}$$

Then the derivative would be

$$\frac{dF(T)}{dT} = \frac{-b - 2cT}{(1 + a + bT + cT^2)^2}$$

- Thomas; The explicit equation is

$$T(T) = ae^{bT} - (c + de^{eT})$$

Then the derivative would be

$$\frac{dT(T)}{dT} = abe^{bT} - dee^{eT}$$

- Quadratic; The explicit equation is

$$Q(T) = a + bT + cT^2$$

Then the derivative would be

$$\frac{dQ(T)}{dT} = b + 2cT$$

- Briere2; The explicit equation is

$$B(T) = aT(T - T_{min})(T_{max} - T)^{\frac{1}{b}}$$

Then the derivative would be

$$\frac{dB(T)}{dT} = a(T - T_{min})(T_{max} - T)^{\frac{1}{b}} + aT(T_{max} - T)^{\frac{1}{b}} + \frac{-aT}{b}(T - T_{min})(T_{max} - T)^{\frac{1-b}{b}}$$

- Johnson Lewin; The explicit equation is

### The partial derivative of each parameter

Now, let calculate the partial derivative for each parameter function by assuming all other parameters are constant

- Transmission rate in human $\beta_h(T)$. The curve was modeled with briere2 function, thus

$$\begin{aligned}\frac{d\mathcal{R}_0}{dT} &= \frac{\partial \mathcal{R}_0}{\partial \beta_h(T)} \frac{\partial \beta_h(T)}{dT} = \frac{\mathcal{R}_0}{2\beta_h(T)} \frac{\partial \beta_h(T)}{dT} \\ &= \frac{\mathcal{R}_0}{2} * \left(\frac{1}{T} + \frac{1}{T - T_{min}} - \frac{1}{b(T_{max} - T)}\right)\end{aligned}$$

- Transmission rate in snail $\beta_s(T)$. The curve was modeled with spain function, thus

$$\begin{aligned}\frac{d\mathcal{R}_0}{dT} &= \frac{\partial \mathcal{R}_0}{\partial \beta_s(T)} \frac{\partial \beta_s(T)}{dT} = \frac{\mathcal{R}_0}{2\beta_s(T)} \frac{\partial \beta_s(T)}{dT} \\ &= \frac{\mathcal{R}_0}{2r_0 e^{aT}(1 - be^{cT})} r_0 e^{aT}(a - abe^{cT} - bce^{cT}) \\ &= \frac{\mathcal{R}_0(a - abe^{cT} - bce^{cT})}{2(1 - be^{cT})}\end{aligned}$$

- Miracidia hatching rate $\delta_e(T)$, The curve was modeled with flinn function, thus

$$\begin{aligned}\frac{d\mathcal{R}_0}{dT} &= \frac{\partial \mathcal{R}_0}{\partial \delta_e(T)}\frac{\partial \delta_e(T)}{dT} = \frac{\mathcal{R}_0}{2\delta_e(T)}\frac{\partial \delta_e(T)}{dT} \\ &= \frac{\mathcal{R}_0}{2\delta_e(T)}\frac{-b-2cT}{(1+a+bT+cT^2)^2} \\ &= \frac{-\mathcal{R}_0(b+2cT)}{2(1+a+bT+cT^2)}\end{aligned}$$

- Cercarial releasing rate $\nu_c(T)$, The curve was modeled with Gaussian function, thus

$$\begin{aligned}\frac{d\mathcal{R}_0}{dT} &= \frac{\partial \mathcal{R}_0}{\partial \nu_c(T)}\frac{\partial \nu_c(T)}{dT} = \frac{\mathcal{R}_0}{2\nu_c(T)}\frac{\partial \nu_c(T)}{dT} \\ &= \frac{\mathcal{R}_0}{2\nu_c(T)}\frac{-r_{max}\left(T-T_{opt}\right)}{a^2}e^{-0.5\left(\frac{T-T_{opt}}{a}\right)^2} \\ &= \frac{-\mathcal{R}_0\left(T-T_{opt}\right)}{2a^2}\end{aligned}$$

- Prepatent period $\sigma_s(T)$, The curve was modeled with Gaussian function, thus

$$\begin{aligned}\frac{d\mathcal{R}_0}{dT} &= \frac{\partial \mathcal{R}_0}{\partial \sigma_s(T)}\frac{\partial \sigma_s(T)}{dT} \\ &= \frac{\mathcal{R}_0}{2}\left(\frac{1}{\sigma_s(T)}-\frac{1}{\left(\sigma_s(T)+\mu_i(T)\right)}\right)\frac{-r_{max}\left(T-T_{opt}\right)}{a^2}e^{-0.5\left(\frac{T-T_{opt}}{a}\right)^2}\end{aligned}$$

- Mortality rate of miracidia, $\mu_m(T)$, The curve was modeled with Thomas function, thus

$$\begin{aligned}\frac{d\mathcal{R}_0}{dT} &= \frac{\partial \mathcal{R}_0}{\partial \mu_m(T)}\frac{\partial \mu_m(T)}{dT} = -\frac{\mathcal{R}_0}{2\mu_m(T)}\frac{\partial \mu_m(T)}{dT} \\ &= -\frac{\mathcal{R}_0}{2\mu_m(T)}(abe^{bT}-dee^{eT}) \\ &= \frac{\mathcal{R}_0(-abe^{bT}+dee^{eT})}{2\left(ae^{bT}-(c+de^{eT})\right)}\end{aligned}$$

- Mortality rate of cercaria, $\mu_c(T)$, The curve was modeled with Spain function, thus

$$\begin{aligned}\frac{d\mathcal{R}_0}{dT} &= \frac{\partial \mathcal{R}_0}{\partial \mu_c(T)}\frac{\partial \mu_c(T)}{dT} = -\frac{\mathcal{R}_0}{2\mu_c(T)}\frac{\partial \mu_c(T)}{dT} \\ &= -\frac{\mathcal{R}_0}{2r_0e^{aT}(1-be^{cT})}r_0e^{aT}(a-abe^{cT}-bce^{cT}) \\ &= -\frac{\mathcal{R}_0(a-abe^{cT}-bce^{cT})}{2(1-be^{cT})}\end{aligned}$$

- Mortality rate of infected snails, $\mu_i(T)$, The curve was modeled with flinn function, thus

$$\begin{aligned}\frac{d\mathcal{R}_0}{dT} &= \frac{\partial\mathcal{R}_0}{\partial\mu_i(T)}\frac{\partial\mu_i(T)}{dT} = \frac{\mathcal{R}_0}{2}\frac{-\big(\sigma_s(T)+2\mu_i(T)\big)}{\mu_i(T)\big(\sigma_s(T)+\mu_i(T)\big)}\frac{\partial\mu_i(T)}{dT} \\ &= \frac{\mathcal{R}_0}{2}\frac{-\big(\sigma_s(T)+2\mu_i(T)\big)}{\mu_i(T)\big(\sigma_s(T)+\mu_i(T)\big)}\frac{-b-2cT}{(1+a+bT+cT^2)^2} \\ &= \frac{\mathcal{R}_0\big(\sigma_s(T)+2\mu_i(T)\big)(b+2cT)}{2\mu_i(T)\big(\sigma_s(T)+\mu_i(T)\big)(1+a+bT+cT^2)^2}\end{aligned}$$
